# Distinct Spatiotemporal Patterns of White Matter Hyperintensity Progression

**DOI:** 10.1101/2025.02.26.25322929

**Authors:** Jinyong Chung, Gilsoon Park, Wi-Sun Ryu, Dawid Schellingerhout, Hang-Rai Kim, Dong Seok Gwak, Elizabeth Haddad, Neda Jahanshad, Beom Joon Kim, Keun-Sik Hong, Hyerin Oh, Sang-Wuk Jeong, Joon-Tae Kim, Man Seok Park, Kang-Ho Choi, Kyungbok Lee, Tai Hwan Park, Sang-Soon Park, Jong-Moo Park, Kyusik Kang, Kyung-Ho Yu, Mi Sun Oh, Soo Joo Lee, Jae Guk Kim, Jae-Kwan Cha, Dae-Hyun Kim, Jun Lee, Moon-Ku Han, Yong-Jin Cho, Byung-Chul Lee, Hee-Joon Bae, Hosung Kim, Dong-Eog Kim

## Abstract

White matter hyperintensity (WMH), a key imaging marker for brain health and the most prevalent brain abnormality in the general population, has prognostic implications for stroke. However, as a prognostic factor, WMH is hard to apply because it varies in both location and size. Reducing this complexity into a set of recognizable patterns would improve the utility of WMH as a cerebrovascular biomarker. In this paper, we show three distinct spatiotemporal patterns of WMH progression in ischemic stroke patients. We used a multicenter MRI database of 9,719 consecutive patients with acute ischemic stroke, plus the UK Biobank database (*n* = 36,210 controls). Subtype and Stage Inference (SuStaIn) modeling was performed to simultaneously subtype and stage WMH based on longitudinal inference from the Korean multicenter cross-sectional data, after normalizing WMH volumes of our stroke patients to the age-adjusted WMH volumes of low-risk controls (*n* = 13,811) from the UK Biobank database. Then, we rigorously characterized demographic profiles, vascular risk factors, and stroke outcomes across three different WMH progression trajectories: fronto-parietal (subtype 1: T1), radial (T2), and temporo-occipital (T3) progression. Subtypes remained consistent between baseline and follow-up (median [interquartile range] years of follow-up: 4.5 [2.8□7.3]) in the majority (> 77%) of patients during WMH progression. T1 showed relatively delayed WMH onset and was enriched for women and hypertension, whereas T3 showed earlier WMH onset and was enriched for men, atrial fibrillation, and coronary artery disease. Hypertension and diabetes mellitus were major risk factors that could accelerate WMH progression in all subtypes. The index stroke, i.e., acute symptomatic infarction, was more likely to be due to small vessel occlusion in T1 and cardioembolism in T3. Regarding post-stroke outcomes, early (< 3 weeks) neurological deterioration by symptomatic hemorrhagic transformation and 3-month unfavorable functional outcome were more frequent in T3, while 1-year ischemic stroke recurrence was more frequent in T1. We also found that our WMH subtyping–staging model and the widely used Fazekas WMH scale were consistent while also offering complementary strengths in profiling stroke risk factors and outcomes. Using our model, high-risk controls (without a history of neurological diseases but with vascular risk factors, *n* = 22,399) in the UK Biobank were assigned to their most likely WMH subtype and stage, demonstrating spatiotemporal patterns of WMH progression that closely paralleled those observed in stroke patients. Of note, we observed distinct stage distributions between high-risk controls and stroke patients, along with a higher area under the curve in the receiver operating curve analysis for distinguishing stroke patients from high-risk controls, compared to that of WMH volume. In conclusion, we identified distinct spatiotemporal trajectories of WMH progression. This WMH subtyping–staging model can capture demographic and vascular risk factors and may provide useful predictions for stroke outcomes across different WMH subtypes and stages. Further prospective investigation is required to confirm whether our model can predict future stroke occurrence more effectively than WMH volume.

**Short abstract:** White matter hyperintensity (WMH), a key imaging biomarker for brain health, is common in the general population and has prognostic implications for stroke. Using a multicenter MRI dataset of 9,719 stroke patients plus the UK Biobank (*n* = 36,210 low- and high-risk controls), we employed Subtype and Stage Inference (SuStaIn) modeling and identified three distinct WMH progression subtypes: fronto-parietal (T1), radial (T2), and temporo-occipital (T3). Longitudinal validation confirmed that this classification was stable in the majority of cases. There were three distinct profiles: i) T1 showed delayed WMH onset and more hypertension, while T3 had more atrial fibrillation and coronary heart disease; ii) T1 and T2 were linked to small vessel occlusion, while T3 was linked to cardioembolism; iii) T1 had higher 1-year ischemic recurrence, while T3 showed a higher incidence of early (< 3 weeks) neurological deterioration by symptomatic hemorrhagic transformation and poorer 3-month outcomes. These results were consistent with and complement total or regional Fazekas scale-related findings. We also observed that i) the spatiotemporal patterns of WMH progression in high-risk controls closely paralleled those observed in stroke patients, and ii) distinct stage distributions between the groups enabled greater discriminatory power, compared to crude WMH volume measures, for distinguishing stroke patients from high-risk controls. In conclusion, this new WMH subtyping–staging model can reliably capture WMH progression, demographic profiles, and vascular risk factors, offering improved predictive power for post-stroke outcomes while showing a potential to predict stroke occurrence.

## Introduction

White matter hyperintensities (WMHs) are among the most common brain abnormalities observed on brain MRI and serve as key imaging biomarkers of brain health. They are typically linked to cerebral small vessel disease (SVD) due to arteriolosclerosis, lipohyalinosis, or other chronic vascular alterations, particularly in elderly individuals with or without vascular risk factors such as hypertension and diabetes^1–11^. WMH burden is a risk factor for stroke and poor functional outcomes following stroke^12–21^. However, WMH distribution varies in both location and size^22,23^, potentially reflecting its diverse underlying pathologies.^24–29^ This complexity limits the utility of WMH as a cerebrovascular biomarker. Identifying distinct spatial subtypes of WMH may contribute to more tailored approaches in stroke prevention and treatment.

The most widely used spatial classification was first introduced by Fazekas et al., 1987^30^. It divides WMH into periventricular white matter region (PVWM) and deep white matter region (DWM), grading their progression independently by using semi-quantitative visual rating scales. Recent approaches have employed more detailed spatial subdivisions based on functional lobes^23,31,32^ or arterial territories^23,33^, along with volumetric measurements. Yet these methods assess WMH progression using only cross-sectional approaches, without considering longitudinal changes^34–36^.

Recently, Subtype and Stage Inference (SuStaIn) modeling^37^, a machine learning-based probabilistic algorithm, has been proposed to identify distinct spatiotemporal trajectories of disease progression by applying longitudinal inference to large cross-sectional datasets. The algorithm assumes that disease progression follows distinct subtypes with subtype-specific stages and that each individual’s phenotype reflects a snapshot of progression at a particular stage for a given subtype. This method has successfully revealed distinct disease progression trajectories for cortical atrophy, tau pathology, brain lesion volume, and T1/T2 ratio, reflecting unique genotypes, demographics, cognitive profiles, and prognoses in various types of dementia^37,38^ and multiple sclerosis^39^. To date, this method has not been used to explore trajectories of WMH progression.

In this study, we employed SuStaIn modeling to identify distinct spatiotemporal trajectories of WMH progression in 9,179 consecutive patients admitted with acute ischemic stroke to 11 academic stroke centers in Korea. We found that, indeed, subtyping WMH severity and location defined distinct spatiotemporal patterns of WMH progression, identified subtypes enriched for certain demographics and vascular risk factors, and proved useful for predicting distinct stroke outcomes.

## Results

### Three distinct spatiotemporal trajectories of WMH progression: fronto-parietal, radial, and temporo-occipital lesion growth

We used SuStaIn modeling^37^ to identify distinct spatiotemporal patterns of WMH progression in 9,179 (5,395 males, 59%) consecutive patients with acute ischemic stroke (Supplementary Fig. 1; Korean MRI-based stroke database^11,40–42^). SuStaIn modeling simulates disease progression as a series of discrete, probabilistic events with a disease-related biomarker reaching predefined levels classified as abnormal^37^. First, we utilized FLAIR MRIs of low-risk controls (*n* = 13,811, UK Biobank) without vascular risk factors to establish baseline distributions of WMH volumes in 20 regions of interest (ROIs) across five functional brain lobes (Supplementary Fig. 2). Next, we assessed WMH severity in stroke patients and high-risk controls (*n* = 22,399, UK Biobank) by normalizing each group’s WMH volumes to the age-adjusted WMH volumes of low-risk controls. Severity can reach a threshold, predefined as a z-score of 1.5 or greater, in each of the 20 ROIs. Therefore, there were 20 total WMH progression stages spanning all 20 ROIs.

Based on a cross-validation (CV) process (Supplementary Fig. 3a), we found that a three-subtype model was optimal for addressing the spatial heterogeneity of WMH progression. Although models with more subtypes showed higher log likelihoods and lower CV-based information criteria (CVIC), indicating better model fits, such models began to plateau beyond three subtypes. Models with more than three subtypes also showed a substantial decrease in progression pattern similarity between CV folds within the same subtypes, signifying a decline in the robustness of detecting subtype-specific progression patterns. Moreover, pattern similarity substantially increased between different subtypes within CV folds, a result that signals poor spatiotemporal discrimination between progression patterns of different subtypes.

The three-subtype model identified distinct subtypes of spatiotemporal WMH progression (Fig. 1a) across five different brain lobes (Supplementary Fig. 2). WMH growth was primarily from the frontal lobe to the parietal lobe in Subtype 1 (T1), radial across all lobes in Subtype 2 (T2), and from the temporal lobe to the occipital lobe in Subtype 3 (T3). In all three subtypes, WMH grew outward from the periventricular regions. These findings were corroborated by voxel-wise frequency mapping of WMH (Supplementary Fig. 4).

**Fig. 1.**
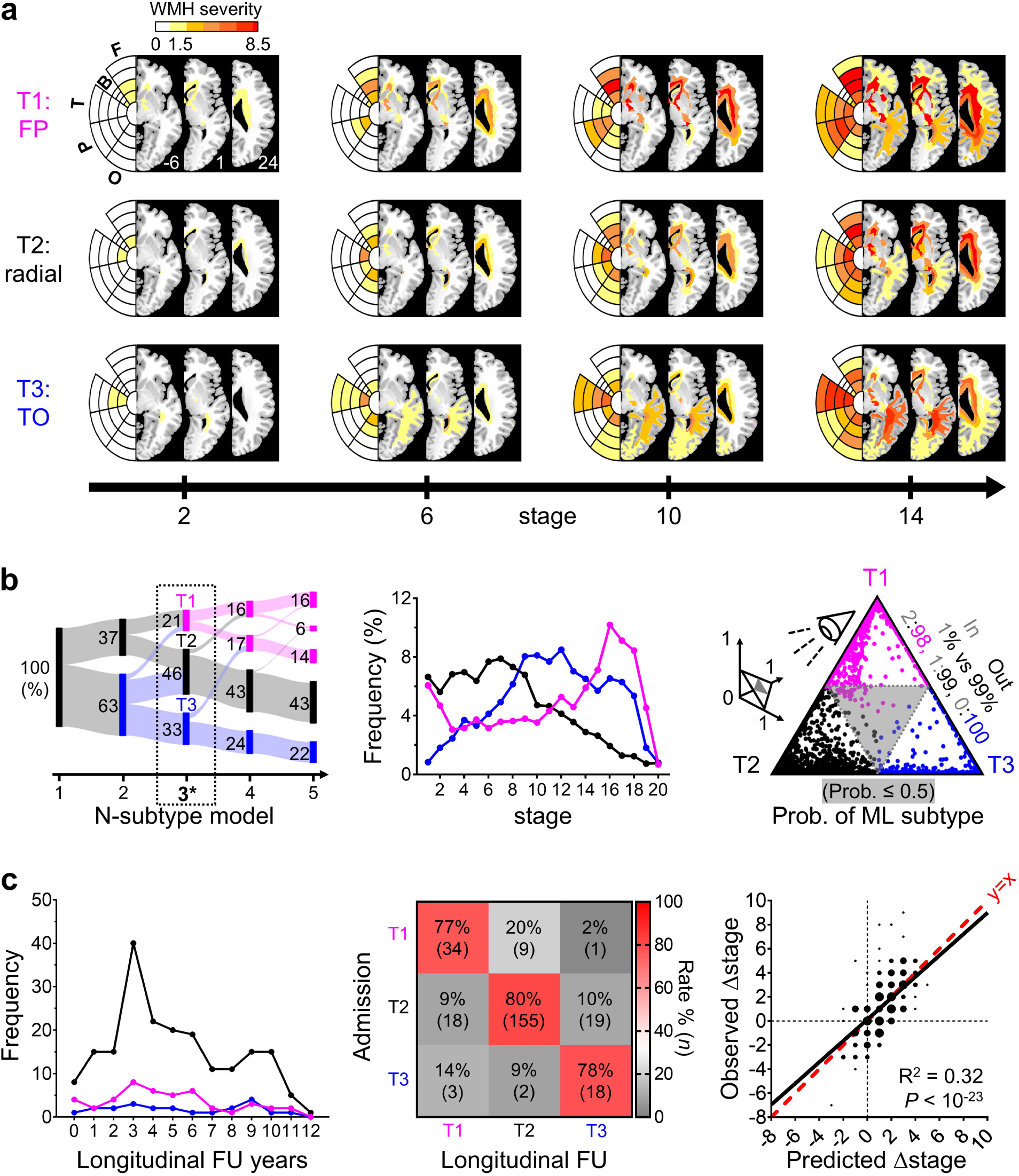
Distinct spatiotemporal trajectories of white matter hyperintensity (WMH) progression in stroke patients. **a**, Spatiotemporal patterns of distinct WMH progression subtypes (T1: fronto-parietal [fp]; T2: radial; T3: temporo-occipital [TO]). Bullseye presentations (F: frontal; B: basal ganglia; T: temporal; P: parietal; O: occipital) and median WMH severity maps are presented for each WMH subtype at WMH stages 2, 6, 10, and 14. The z-axis coordinates in the Montreal Neurological Institute space appear in the bottom right corner of the first three brain slices in the T1 group. **b**, Left, Sankey diagram illustrating patient distributions across the number of subtypes in Subtype and Stage Inference (SuStaIn) modeling. Main results in this study utilized the three-subtype model (*). Groups in models were assigned the same color as the subtype with the highest patient proportion based on the three-subtype model. Middle, stage distributions across subtypes. Patients at stage 0 were not included in any other subtypes (T1∼T3). Right, scatter plot showing maximum likelihood (ML) subtype probabilities (prob.) of individuals, in a 2D projection on a triangular plane. **c**, Left, distributions of follow-up (FU) years across subtypes. Middle, longitudinal subtype stability rates (number of patients) by subtypes at admission and FU. Right, simple linear regression graph of observed vs. predicted stage changes in stroke patients after FU. Scatter dot sizes represent number of patients (*n* = 1, 2 ≤ *n* ≤ 5, 6 ≤ *n* ≤ 10, and 11 ≤ *n* ≤ 17). These findings suggest baseline assignment to a subtype is mostly stable, with relatively few patients crossing out of their initially assigned group.

Of all patients (*n* = 9,179), 443 (5%) were assigned as stage 0 and were not classified to any subtype (T1∼T3). Among the remaining 8,736 patients, 1,870 (21%), 4,014 (46%), and 2,852 (33%) were classified as T1, T2, and T3, respectively. In addition to these three-subtype model data, the Sankey diagram (Fig. 1b, left) shows how the proportional distribution of each subtype changes as the number of subtypes expands, ranging from 1 to 5. The selected three-subtype model effectively captured the full progression of WMH, as patients were distributed across all 20 stages within each subtype (Fig. 1b, middle). Of note, patient distribution across WMH stages differed among WMH subtypes. Thus, WMH stage was included as a covariate in subsequent analyses to compare demographics, risk factors, stroke etiology, stroke severity, and post-stroke outcomes across WMH subtypes. As shown in the probability of maximum likelihood (ML) subtype plot (Fig. 1b, right; a 2D projection on a triangular plane) presenting each patient’s probability of being classified as the most likely subtype, approximately 99% (*n* = 8,673) of all patients showed good fits to the model (ML subtype probability ≥ 0.5, outside of the gray-shaded triangle).

We also determined whether an assigned subtype category remained consistent through stage advancement of WMH progression over time. Indeed, we assessed this subtype stability to validate the longitudinal robustness of classifying distinct WMH growth trajectories in individual patients. To evaluate this, we used a longitudinal dataset of 264 patients’ fluid-attenuated inversion recovery (FLAIR) MRI at admission and follow-up (FU) (median [interquartile range; IQR] years: 4.5 [2.8□7.3]; Fig. 1c, left). Five patients (2%) were assigned to stage 0 at admission and were excluded from the analysis. In the majority (> 77%) of the 259 included patients, subtypes at admission did not change at FU (Fig. 1c, middle), thereby indicating subtype categorization is consistent over time. This subtype stability was not related to FU period duration (Supplementary Table 1, odds ratio [OR] and 95% confidence interval [CI]: 0.99 [0.89–1.11]). Notably, patients with diabetes mellitus (OR [95% CI]: 0.43 [0.22–0.82]) or those who underwent revascularization therapy (OR [95% CI]: 0.27 [0.08–0.87]) had lower subtype stability at FU when compared to patients without diabetes mellitus or revascularization therapy. Next, we predicted stage progression from admission to FU by employing a mixed-effects regression model that incorporated demographic and vascular risk factors, FU years, and WMH subtype and stage at admission (Fig. 1c, right). Through leave-one-out CV, the predicted stage progressions moderately correlated with observed stage progressions (R^2^ > 0.3, *P* < 10^−23^).

### Distinct demographic and risk factor profiles in different WMH subtypes with relatively delayed WMH onset, more women, and more hypertension in T1 vs. earlier WMH onset, more males, and more atrial fibrillation (AF) in T3

Table 1 shows baseline characteristics of patients with different WMH progression subtypes. Fig. 2 presents raw means (scatter dots) and estimated marginal means (EMM; a curved line with shaded 95% CI, obtained from mixed-effects regression models after adjusting for age and sex) along the stage in each subtype. Detailed statistical information is presented in Supplementary Table 2. If the *P* for the subtype-stage interaction was less than 0.1, we conducted subgroup analyses by dividing patients into two groups, namely those with stage ≤ 9 (median) and those with stage > 9, to assess how subtype differences varied across stages.

**Fig. 2.**
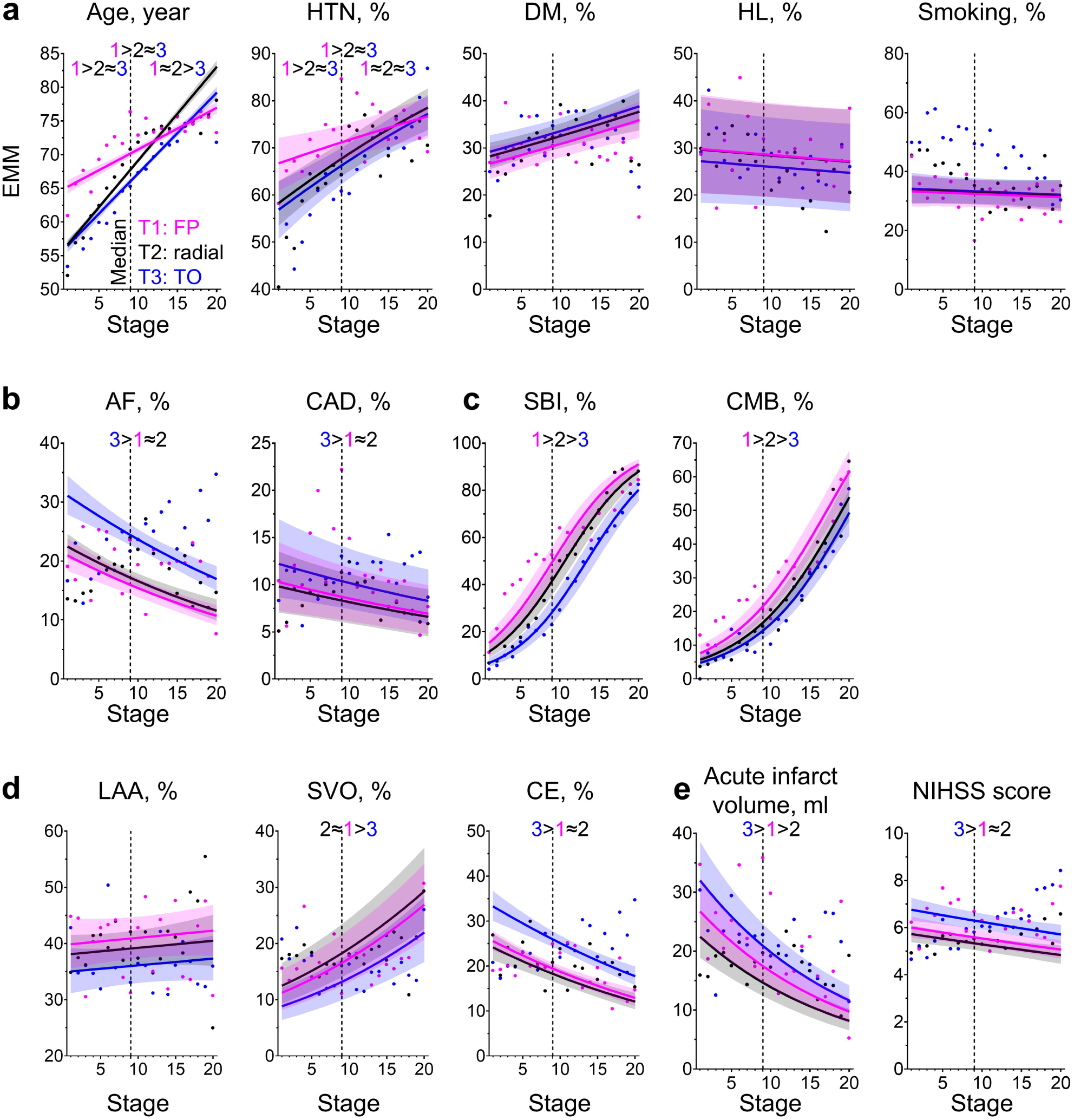
Distinct demographics, risk factor, and etiology profiles of stroke patients with different white matter hyperintensity (WMH) subtypes. Graphs of estimated marginal means (EMMs; curved line with shaded 95% confidence interval) for demographics, vascular risk factors, and etiologies across WMH stages within each WMH subtype (T1: fronto-parietal [fp]; T2: radial; T3: temporo-occipital [TO]) in stroke patients. Scatter plots of raw means were superimposed. The graph explicitly presents significant inter-subtype differences across all stages. For factors showing significant subtype–stage interactions, we further stratified stages into early and late periods based on median stage (9 for stroke patients) in order to derive inter-subtype differences across these periods. **a,** Age, hypertension (HTN), diabetes mellitus (DM), hyperlipidemia (HL), and current smoking. **b,** Atrial fibrillation (AF) and coronary artery disease (CAD). **c**, Presence of silent brain infarcts (SBIs) and cerebral microbleeds (CMBs). **d**, Stroke subtypes: large artery atherosclerosis (LAA), small vessel occlusion (SVO), and cardioembolism (CE). **e**, Acute infarct volume and admission National Institute of Health Stroke Scale (NIHSS) score.

**Table 1.** Demographics, vascular risk factors, and stroke etiologies by subtype and stage of white matter hyperintensity progression in stroke patients.

|  | **T1: FP** | **T2: radial** | **T3: TO** | ***P*\*** |  |  |
| --- | --- | --- | --- | --- | --- | --- |
| ***n* (% of total)** | 1870 (21.4) | 4014 (46.0) | 2852 (32.7) | **subtype** | **stage** | **interaction** |
| **Age, year (SD)** | 72.4 (10.8) | 66.2 (13.1) | 67.9 (12.0) | < 0.001 | < 0.001 | < 0.001 |
| **Sex, male %** | 46.7 | 54.7 | 72.3 | < 0.001 | < 0.001 | - |
| **HTN, %** | 73.2 | 62.7 | 65.3 | 0.007 | < 0.001 | 0.091 |
| **DM, %** | 30.1 | 29.8 | 32.8 | 0.167 | < 0.001 | - |
| **HL, %** | 28.6 | 27.8 | 26.5 | 0.104 | 0.260 | - |
| **Smoking, %** | 31.8 | 38.9 | 47.6 | 0.879 | 0.453 | - |
| **AF, %** | 19.8 | 18.5 | 24.2 | < 0.001 | < 0.001 | - |
| **CAD, %** | 10.7 | 8.9 | 11.4 | 0.017 | 0.007 | - |
| **Presence of SBI, %** | 58.4 | 34.2 | 40.1 | < 0.001 | < 0.001 | - |
| **Presence of CMB, %** | 30.0 | 14.8 | 20.4 | < 0.001 | < 0.001 | - |

| **Stroke subtypes** |  |  |  |  |  |  |
| --- | --- | --- | --- | --- | --- | --- |
| LAA, % | 38.6 | 38.5 | 36.7 | 0.071 | 0.269 | - |
| SVO, % | 16.7 | 17.5 | 15.0 | < 0.001 | < 0.001 | - |
| CE, % | 20.7 | 20.3 | 24.5 | < 0.001 | < 0.001 | - |
| **Acute infarct volume (IQR), ml** | 2.46<br>(0.64–13.77) | 2.36<br>(0.57–13.14) | 3.53<br>(0.60–18.21) | < 0.001 | < 0.001 | - |
| **Admission NIHSS score (IQR)** | 4 (2–9) | 3 (1–8) | 4 (2–9) | < 0.001 | < 0.001 | - |

\**P* from mixed-effects regression models that included each variable along with age (except in the case of age), sex (except in the case of sex), WMH subtype, WMH stage, and subtype–stage interaction as fixed effects and imaging center as a random effect. Onset to MRI time was added as a fixed factor for acute infarct volume. FP, fronto-parietal; TO, temporo-occipital; SD, standard deviation; HTN, hypertension; DM, diabetes mellitus; HL, hyperlipidemia; AF, atrial fibrillation; CAD, coronary artery disease; SBI, silent brain infarct; CMB, cerebral microbleeds; LAA, large artery atherosclerosis; SVO, small vessel occlusion; CE, cardioembolism; IQR, interquartile range; NIHSS, National Institute of Health Stroke Scale.

Mean age showed significant interactions with subtype and stage (Table 1; *P* < 0.001), although age increased as WMH stage advanced in all subtypes (Fig. 2a and Supplementary Table 2). At earlier stages (≤ 9), the mean age was higher in T1, a pattern that indicated relatively delayed WMH onset in this subtype compared to T2 and T3 (Supplementary Table 3). At later stages (> 9), mean ages were similar between T1 and T2 but still lower in T3 (Supplementary Table 4). As shown in Table 1, the proportion of men was lower in T1 (46.7%) and T2 (54.7%) than in T3 (72.3%; both *P* < 0.001).

After adjusting for age and sex, hypertension was most frequently observed in T1 (at earlier stages), while AF was most frequently observed in T3 (Fig. 2a, b). Hypertension showed a trend towards rising prevalence in all subtypes, particularly at earlier stages (≤ 9), and subsequently reached a plateau without significant inter-subtype differences in frequency (Supplementary Tables 3 and 4). AF exhibited a decreasing trend across all stages in all subtypes. These findings suggest that patients with AF are less likely to remain stroke-free until they develop severe WMH, compared to those without AF. Our findings were similar for coronary artery disease. There was no significant inter-subtype difference in the prevalence of diabetes mellitus, hyperlipidemia, or current smoking. Of note, only diabetes mellitus showed an increasing trend with advancing stages in all subtypes. Unadjusted data of demographics and risk factor profiles are shown in Supplementary Fig. 5a-d and Supplementary Table 5.

### Chronic and acute stroke-related lesions have distinct etiological features in different WMH subtypes, with more frequent small vessel occlusion (SVO) in T1 and more frequent cardioembolism (CE) in T3

Silent brain infarcts and cerebral microbleeds as key indicators of SVD^43^ appeared most frequently in T1 and least frequently in T3 (Table 1 and Fig. 2c). In all subgroups, the proportion of patients with chronic lesions (silent brain infarcts and cerebral microbleeds) increased with advancing stages. Detailed statistical information is presented in Supplementary Table 6. In line with the association between T1 and SVD markers, acute infarction in T1 was more likely to be due to SVO (Fig. 2d) compared to that in T3. Reflecting the aforementioned higher prevalence of AF in T3, CE stroke occurred more frequently in T3 across all stages than in other subtypes. Similar to T1, T2 showed a higher proportion of SVO and a lower proportion of CE than T3. The proportion of SVO strokes expanded with progressing WMH stages after adjusting for age and sex. This finding aligned with, as noted above, rising hypertension prevalence as WMH stages advance. After age, hypertension is the most important risk factor for SVD^44^ and WMH^11,45,46^. As in AF, the proportion of CE strokes decreased with progressing WMH stages. Large artery atherosclerotic strokes did not show notable trends with advancing WMH stages. As expected, acute infarct volumes were relatively high in T3 (Fig. 2e). Further, acute infarct volumes diminished as WMH stages advanced in all WMH subtypes with or without adjusting for age and sex (Supplementary Fig. 5e and Supplementary Table 5). Admission National Institute of Health Stroke Scale (NIHSS) scores data appeared to corroborate infarct volume data when adjusted for age and sex (Fig. 2e) but without such adjustment, NIHSS scores showed an increasing trend as WMH stages advanced (Supplementary Fig. 5e and Supplementary Table 5).

### Different WMH subtypes have distinct stroke outcomes: T3 exhibits more frequent early symptomatic hemorrhagic transformation and 3-month unfavorable functional outcome while T1 has more frequent 1-year ischemic stroke recurrence

We compared 3-week (in-hospital), 3-month, and 1-year outcomes in the three different WMH progression subtypes after adjusting for age, sex, and revascularization therapy (Table 2 and Fig. 3a-c). Early neurological deterioration within 3 weeks and its most common cause, stroke progression, occurred more frequently with advancing WMH stages in all subtypes, without significant inter-subtype differences (Fig. 3a). Symptomatic hemorrhagic transformation appeared more often in T3 than in other subtypes. Intriguingly, the incidence of symptomatic hemorrhagic transformation tended to decrease with WMH progression. Stroke recurrence did not change as WMH stages advanced, with no significant differences observed within or between subtypes. At 3 months, the proportion of patients with unfavorable functional outcome (modified Rankin Scale [mRS] score > 3) was larger in T3 than in other subtypes (Fig. 3b). Unfavorable functional outcome levels were higher with advancing WMH stages in all subtypes. The 1-year recurrence of ischemic stroke was notably higher in T1 than in other WMH subtypes, regardless of their stages (Fig. 3c and Table 2). However, hemorrhagic stroke recurrence, which was less frequent than ischemic stroke recurrence, expanded with WMH progression in all subtypes, showing no significant inter-subtype difference. All-cause death and nonvascular death within 1 year rose with advancing WMH stages, but these increases did not differ among WMH subtypes. Vascular death was not related to either WMH subtypes or stages (Table 2).

**Fig. 3.**
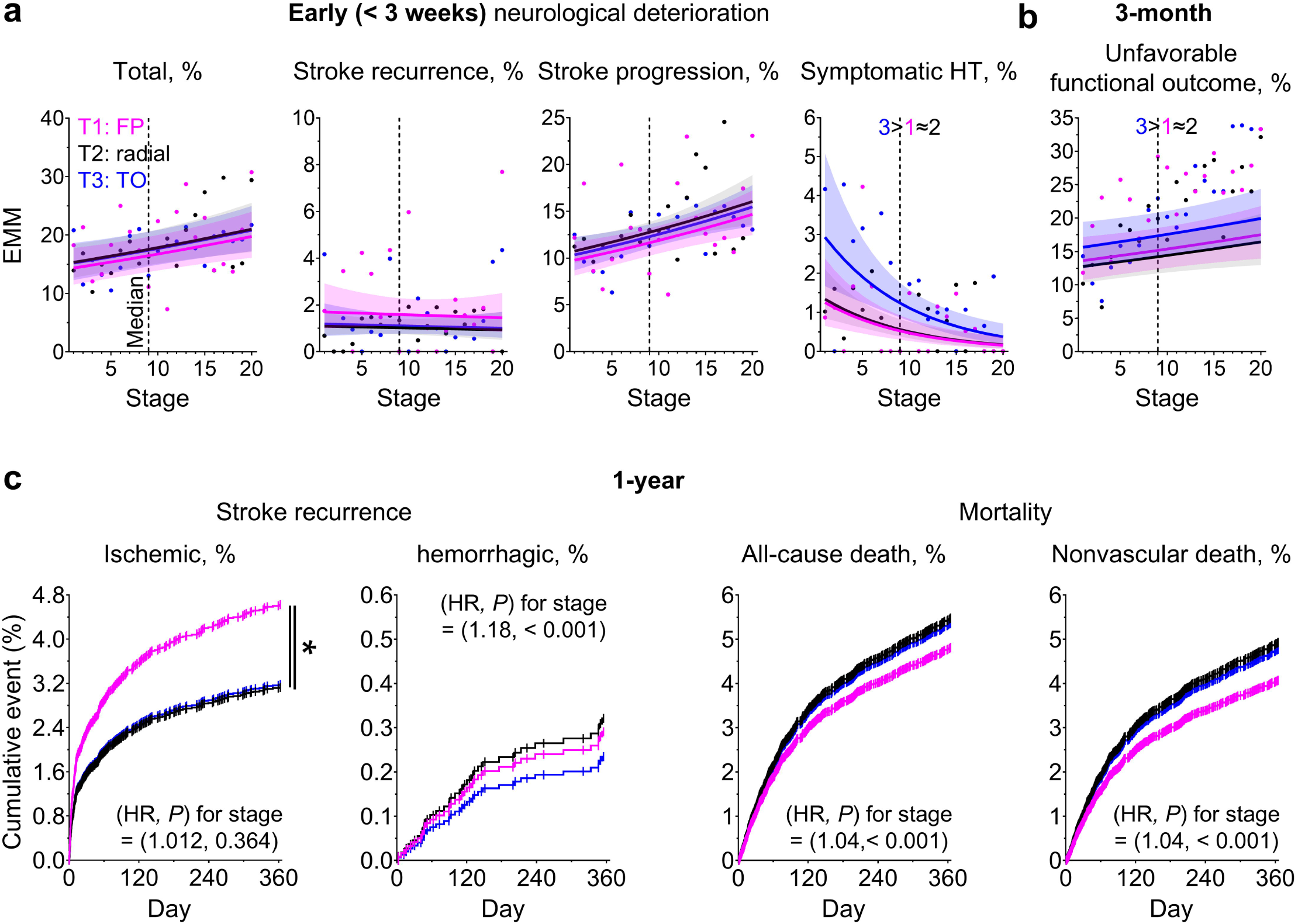
Post-stroke outcomes by subtype and stage of white matter hyperintensity (WMH) progression in stroke patients. Graphs of estimated marginal means (EMMs; curved line with shaded 95% confidence interval) for post-stroke outcomes by WMH stage in each WMH subtype (T1: fronto-parietal [FP]; T2: radial; T3: temporo-occipital [TO]). Scatter plots of raw means were superimposed. **a**, Early (< 3 weeks) neurological deterioration and its causes (stroke recurrence, stroke progression, and symptomatic hemorrhagic transformation [HT]). **b**, Unfavorable functional outcome at 3 months (modified Rankin Scale score > 3). **c**, Stroke recurrence (ischemic and hemorrhagic) and mortality (all-cause and nonvascular death) within 1 year. Hazard ratio (HR) and *P* for stage are explicitly presented. Note that ischemic stroke recurrence and mortalities for T2 (black) and T3 (blue) are very similar, with closely overlapping curves.

**Table 2.**
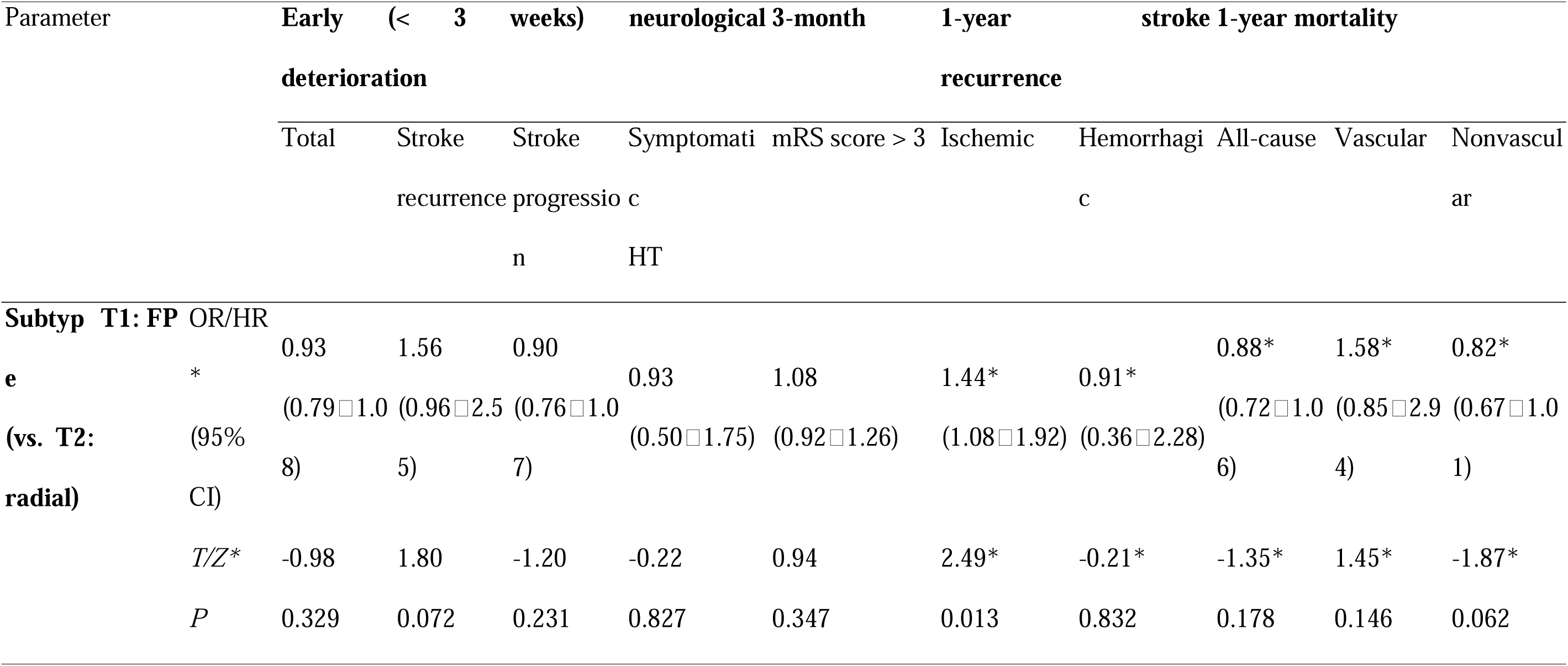
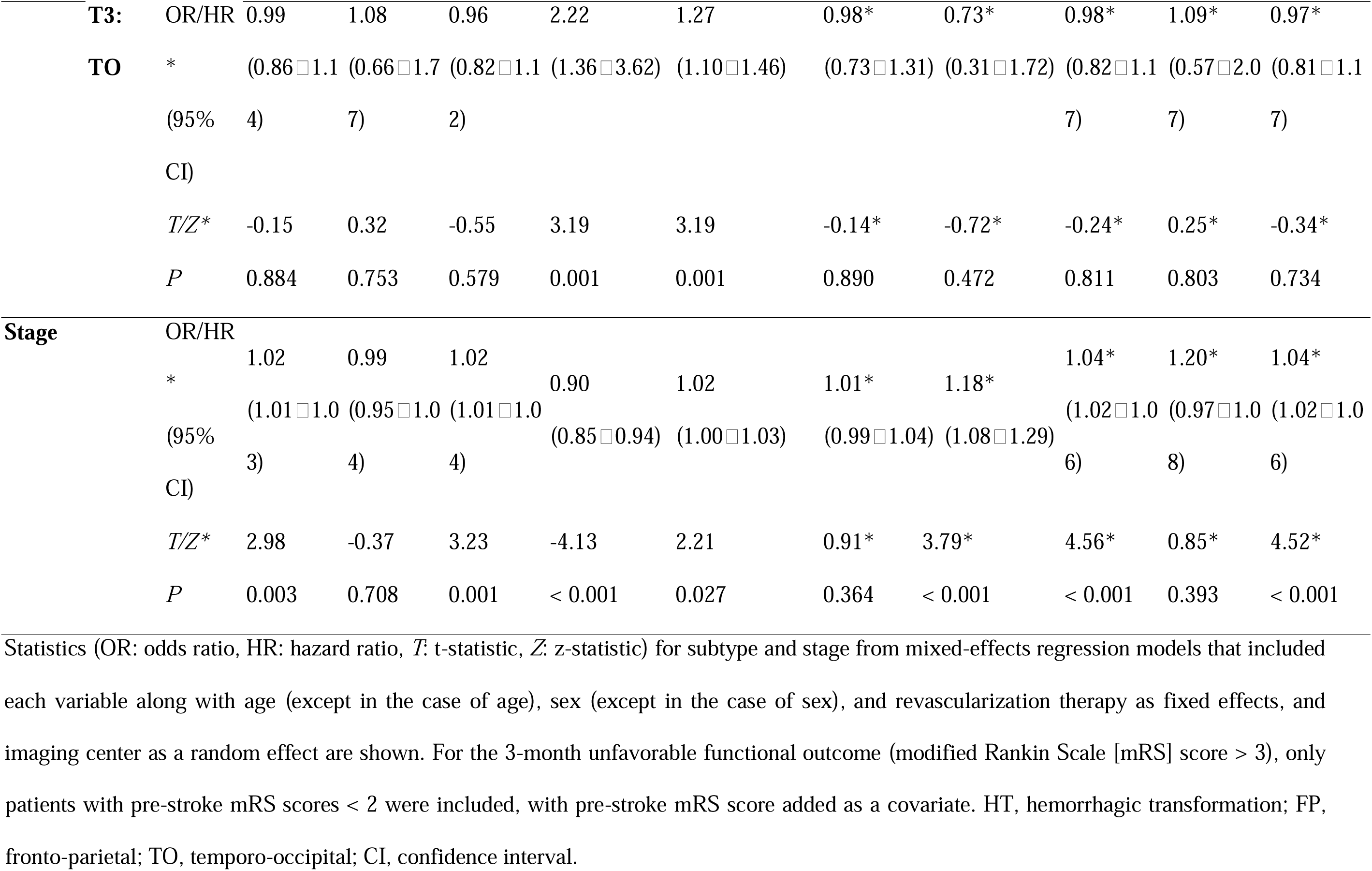
Mixed-effects and cox proportional hazards regression models for post-stroke outcomes according to white matter hyperintensity subtype and stage in stroke patients.

### Alignment and complementarity between the WMH subtyping–staging model and the Fazekas scale in stroke risk and outcomes

To evaluate alignment between the widely used WMH grading system and our proposed WMH subtyping–staging model, we assessed the relationships Fazekas scale scores (PVWMH 0-3, DWMH 0-3, and total 0-6) have with demographics, vascular risk factors, stroke etiology and severity, and post-stroke outcomes. PVWMH and total Fazekas scale scores were associated with age, sex, hypertension, diabetes mellitus, AF, and the presence of silent brain infarcts and cerebral microbleeds (Supplementary Table 7 and 8), patterns that are consistent with connections observed between these vascular risk factors and WMH stages in our model (Supplementary Table 2). DWMH score was associated with age, hypertension, and the presence of silent brain infarcts and cerebral microbleeds. Current smoking and hyperlipidemia were not significantly associated with either Fazekas scale scores or WMH stage in our model. Nevertheless, WMH stage did show a significant relationship with coronary artery disease. As previously mentioned, our model also demonstrated that: i) older and hypertensive patients were more prevalent in T1, and ii) patients with AF and coronary artery disease were more prevalent in T3.

Similar to WMH stage in our model, PVWMH and total Fazekas scale scores were associated with SVO and CE, although DWMH score was associated only with SVO (Supplementary Table 8). Acute infarct volume and admission NIHSS score were associated with PVWMH and total Fazekas scale scores but not with DWMH score. As mentioned above, our model also showed: i) more frequent SVO in T1 and T2 (vs. T3) and more frequent CE in T3 (vs. T1 and T2) and ii) higher acute infarct volumes and admission NIHSS scores in T1 and T3 (vs. T2) and T3 (vs. T2), respectively, as described previously (Supplementary Table 2).

Total Fazekas scale score was associated with early neurological deterioration and its causes, including stroke progression and symptomatic hemorrhagic transformation, but not stroke recurrence within 3 weeks (Supplementary Table 9). However, PVWMH and DWMH scale scores were not significantly associated with early neurological deterioration or its causes except that the PVWMH scale score was negatively associated with symptomatic hemorrhagic transformation. As detailed above, these early neurological deterioration-related findings aligned with WMH stage results in our model, and WMH subtype provided additional predictive information for symptomatic hemorrhagic transformation (Table 2). Unlike our WMH subtypes or stages, total and regional Fazekas scales did not significantly predict unfavorable functional outcome at 3 months. Total Fazekas scale predicted hemorrhagic stroke recurrence (but not ischemic stroke recurrence) and all-cause and nonvascular mortality (but not vascular mortality) within 1 year, whereas regional Fazekas scales predicted none of these outcomes. These 1-year outcome-related findings were, as described previously, consistent with results for our WMH stages. Additionally, our proposed WMH subtypes showed significant inter-subtype differences in 1-year ischemic stroke recurrence.

### Similar spatiotemporal patterns of WMH progression but distinct WMH stage distributions in high-risk controls and stroke patients

Using the three-subtype model we developed by employing data from our stroke patients (vs. data from the low-risk controls in the UK Biobank), we assigned the high-risk controls (without a history of neurological diseases but with vascular risk factors, *n* = 22,399 in the UK Biobank) to their most likely WMH subtype and stage (Fig. 4a). The spatiotemporal patterns of WMH progression, paralleling those observed in stroke patients (Fig. 1a), demonstrated fronto-parietal (T1), radial (T2), and temporo-occipital (T3) WMH progression. After excluding ∼56% of the high-risk controls classified as stage 0, the remaining 9,967 were categorized as T1 (*n* = 558, 46%), T2 (*n* = 4,155, 42%), and T3 (*n* = 1,254, 13%) (Fig. 4b, left). In all subtypes, high-risk controls were predominantly concentrated in the early stages (median [IQR] = 3 [1□6]), and their frequencies were a function of stage following an exponential decrease (Fig. 4b, middle). The stroke population-based three-subtype model effectively classified 86% (*n* = 8,600) of high-risk controls (ML subtype probability ≥ 0.5, outside of the gray-shaded triangle; Fig. 4b, right). As noted previously, the value was 99% for the stroke population itself.

**Fig. 4.**
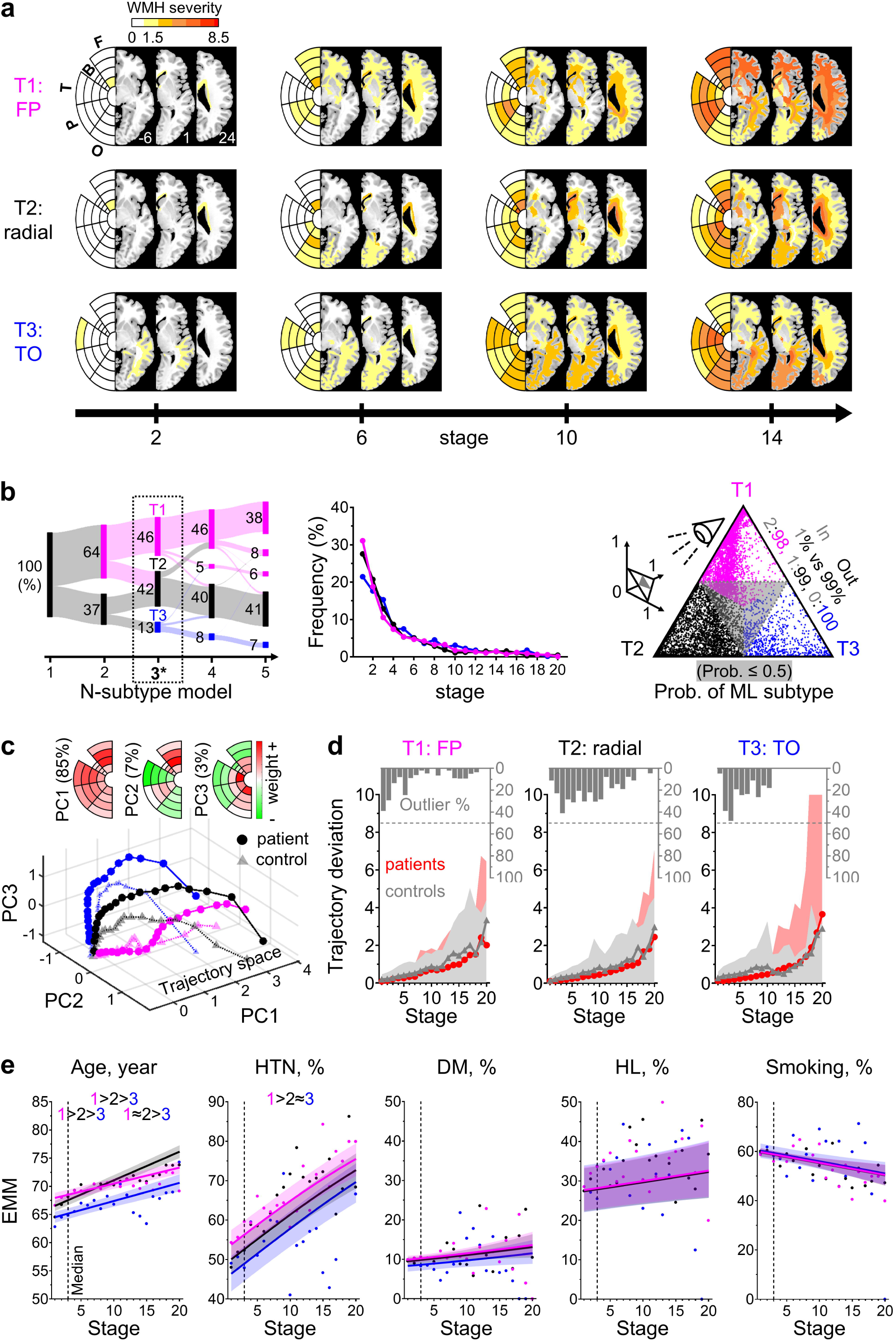
Similar spatiotemporal trajectories of white matter hyperintensity (WMH) progression in high-risk controls (vs. stroke patients). **a**, Spatiotemporal patterns of distinct WMH progression subtypes (T1: fronto-parietal [FP]; T2: radial; T3: temporo-occipital [TO]) in high-risk controls. Bullseye presentations (F: frontal; B: basal ganglia; T: temporal; P: parietal; O: occipital) and median WMH severity maps are presented for each WMH subtype at WMH stages 2, 6, 10, and 14. The z-axis coordinates in the Montreal Neurological Institute space are displayed in the bottom right corner of the first three brain slices in the T1 group. **b**, Left, Sankey diagram illustrating distributions of high-risk controls across the number of subtypes in Subtype and Stage Inference (SuStaIn) modeling. Main results in this study utilized the three-subtype model (*). Groups in models were assigned the same color as the subtype with the highest proportion of patients based on the three-subtype model derived from stroke patients. Middle, distributions of stages across the subtypes. Individuals at stage 0 were not included in any other subtypes (T1∼T3). Right, scatterplot showing maximum likelihood (ML) subtype probabilities (prob.) of individuals, in a 2D projection on a triangular plane. **c**, Spatiotemporal WMH progression in the stroke vs. high-risk control groups, depicted as connected dots (with dense and light coloring, respectively) in the shared representative three-dimensional trajectory space based on the first three principal components (PCs) derived by using the 60 WMH severity maps of the stroke group (Fig. 1a); the maps, which depict median WMH severity for each of the 20 ROIs at 20 stages and three subtypes, were initially expressed as a set of 60 (20 stages × 3 subtypes) coordinates in a 20-dimensional space, where each dimension corresponded to a specific ROI. Each PC was z-normalized using the mean and standard deviation calculated from the combined transformed data of both stroke patients and high-risk controls. Bullseye presentations show the weights (ranging from −0.5 to 0.5) of the three PCs. **d**, Trajectory deviations in spatiotemporal WMH progression between stroke patients and high-risk controls across stages at each subtype, showing the median values (dots) and their 95^th^ percentiles (shaded area) for Euclidean distances from the median of the stroke group. Within-population 95^th^ percentiles for the stroke group are also displayed. Gray bars at the top of the graph represent, among high-risk controls, the proportion of outliers, defined as those with trajectory deviations exceeding the 95^th^ percentile observed in stroke patients for the corresponding subtype and stage. **e**, Graphs of estimated marginal means (EMMs; curved line with a shaded 95% confidence interval) for demographics and vascular risk factor across stages within each subtype in high-risk controls: age, hypertension (HTN), diabetes mellitus (DM), hyperlipidemia (HL), and current smoking. Scatter plots of raw means were superimposed. The graph explicitly presents significant inter-subtype differences across all stages. For factors showing significant subtype–stage interactions, we further stratified the stages into early and late periods based on the median stage (three for high-risk controls), thus defining inter-subtype differences across these periods. See Supplementary Fig. 6 for atrial fibrillation and coronary artery disease.

In addition, spatiotemporal patterns of WMH progression in high-risk controls appeared comparable to those observed in stroke patients, as indicated by quantifying spatial and severity-related differences in WMH between the two respective maps at each stage within each subtype to estimate the degree of alignment in their progression trajectories (Fig. 4c, d). First, we performed principal component analysis (PCA)^47–49^ to identify the principal components (PCs) that account for the greatest variance among the different WMH subtypes and stages in the 60 WMH maps for the stroke group (Fig. 1a). Because the maps represent median WMH severity for each of the 20 ROIs at 20 stages and three subtypes, the data for both the stroke group and the high-risk control group can be represented by a set of 60 (20 stages × 3 subtypes) coordinates in a 20-dimensional space, with each dimension corresponding to a specific ROI. The following PCs, which collectively accounted for 95% of the total variance, were identified from the stroke group data (Fig. 4c, top): PC1 exhibited positive weights for WMH severity across all ROIs, PC2 showed positive weights for fronto-parietal and negative weights for temporo-occipital ROIs, and PC3 displayed positive weights in PVWM (layers 1 and 2) and negative weights in DWM (layers 3 and 4) ROIs.

Next, we used the three leading PCs, as identified by the PCA, to construct a three-dimensional trajectory space (Fig. 4c, bottom). In this space, data from both the stroke group data and the high-risk control group were plotted by transforming the aforementioned 60 coordinates of each group into the trajectory space. The loading weights on the three PCs (Fig. 4c, top) served as the transformation matrices for the 60 original coordinates of the stroke group, reflecting the contribution of each ROI to PC1, PC2, and PC3 in descending order of their explained variance. In the three-dimensional plots (Fig. 4c, bottom), black and blue dots represent the stroke group, while lighter-colored triangles denote the high-risk control group. These analyses revealed similar trajectories for both groups.

Finally, to estimate the trajectory deviation between the stroke group and the high-risk control group in WMH spatiotemporal progression, we calculated the Euclidean distances between the three-dimensional coordinates of individual high-risk controls and the corresponding median value of the stroke group for each stage in each subtype, deriving a total of 60 (20 stages × 3 subtypes) median values and their 95^th^ percentiles. We also calculated within-population median values and 95^th^ percentiles for the stroke patients. As shown in Fig. 4d, the trajectories reflecting the spatiotemporal progression of WMH closely overlapped between the stroke and high-risk control populations.

The high-risk control group and the stroke group exhibited similarities in demographic and risk factor profiles across WMH subtypes. Older age at earlier stages (< 3) and higher proportions of females and hypertension (Fig. 4e and Supplementary Table 10). We observed no significant inter-subtype differences in the prevalence of diabetes mellitus, hyperlipidemia, or current smoking. Trends in both age and the prevalence of hypertension and diabetes mellitus increased with advancing stages, consistent with patterns observed in the stroke group. We also found some dissimilarities between the groups. In the high-risk control group, the proportions of AF and coronary artery disease did not differ significantly across subtypes (Supplementary Fig. 6 and Supplementary Table 10 vs. Fig. 2b and Table 1). Also, the proportions of hyperlipidemia and current smoking significantly changed with advancing stages (Fig. 4e and Supplementary Table 10 vs. Fig. 2a and Table 1).

Given the concordant WMH progression trajectories, it is also noteworthy that WMH stage distributions clearly diverged between high-risk controls and stroke patients (Fig. 1b, 4b and Supplementary Fig. 7a). As mentioned above, the stage distribution of high-risk controls predominantly clustered in the early stages (Fig. 4b, middle), but stroke patients were markedly different (Fig. 1b, middle): T1 presented consistently high frequencies up to stage 9, followed by a linear reduction; T2 showed increasing frequencies up to stage 9, followed by a plateau and then an initially gradual but later sharp decrease; and T3 displayed relatively low frequencies up to stage 14, followed by a late sharp rise and subsequent decline. This clear divergence in WMH stage distributions was further supported by the area under the receiver operating characteristic (ROC) curve, which, compared to WMH volume, demonstrated more pronounced distinctions between stroke patients and high-risk controls (Supplementary Fig. 7b).

### Using cerebral arterial territories provides an alternative but less robust model of spatiotemporal WMH progression

In addition to our proposed model based on ROIs representing frontal, basal ganglia, temporal, parietal, and occipital lobes, we explored an alternative WMH subtyping–staging approach (Fig. 5) based on ROIs representing cerebral arterial territories (anterior, middle, posterior cerebral arteries [ACA, MCA, PCA], and their border zones) and distance from the ventricular surface (Supplementary Fig. 8). SuStaIn modeling and a CV process led to the development of a two-subtype model (Fig. 5a). Models with more than two subtypes showed substantially reduced progression pattern similarity across CV folds, a result that indicates lower model robustness, along with markedly elevated maximum pattern similarity between different subtypes, a result that indicates poorer spatial discrimination between progression patterns of different subtypes (Supplementary Fig. 3b). However, the two-subtype model’s CV evaluation metrics (log likelihood, CVIC, and pattern similarities) also demonstrated poorer model fit, robustness, and spatial discrimination between subtypes than our primary proposed model (Supplementary Fig. 3a). Subtype 1 (T1’) showed major WMH progression starting in the MCA territory and border zones of MCA-PCA and MCA-ACA, while Subtype 2 (T2’) began with major WMH progression in the MCA-PCA border zone and the PCA territory. Nevertheless, these progression patterns were less distinct than those in our proposed model (Fig. 1a). Moreover, these subtypes did not show significant differences in incidence of early (< 3 weeks) neurological deterioration (symptomatic hemorrhagic transformation; linear mixed-effects regression model, *P* for subtype = 0.561), unfavorable functional outcome at 3 months (linear mixed-effects regression model, *P* for subtype = 0.347), or stroke recurrence within 1 year (Cox proportional hazards model, *P* = 0.794 for T2’ [vs. T1’]) (Fig. 5b). Furthermore, in the longitudinal subtype-stability analysis for this two-subtype model, only 67% of patients who exhibited the T2’ subtype at admission remained in the same subtype at FU (Fig. 5c), which is a particularly low rate given that the model includes only two subtypes. Our proposed three-subtype model, by contrast, showed subtype stabilities of more than 77% for all subtypes, despite having one more subtype (Fig. 1c, middle).

**Fig. 5.**
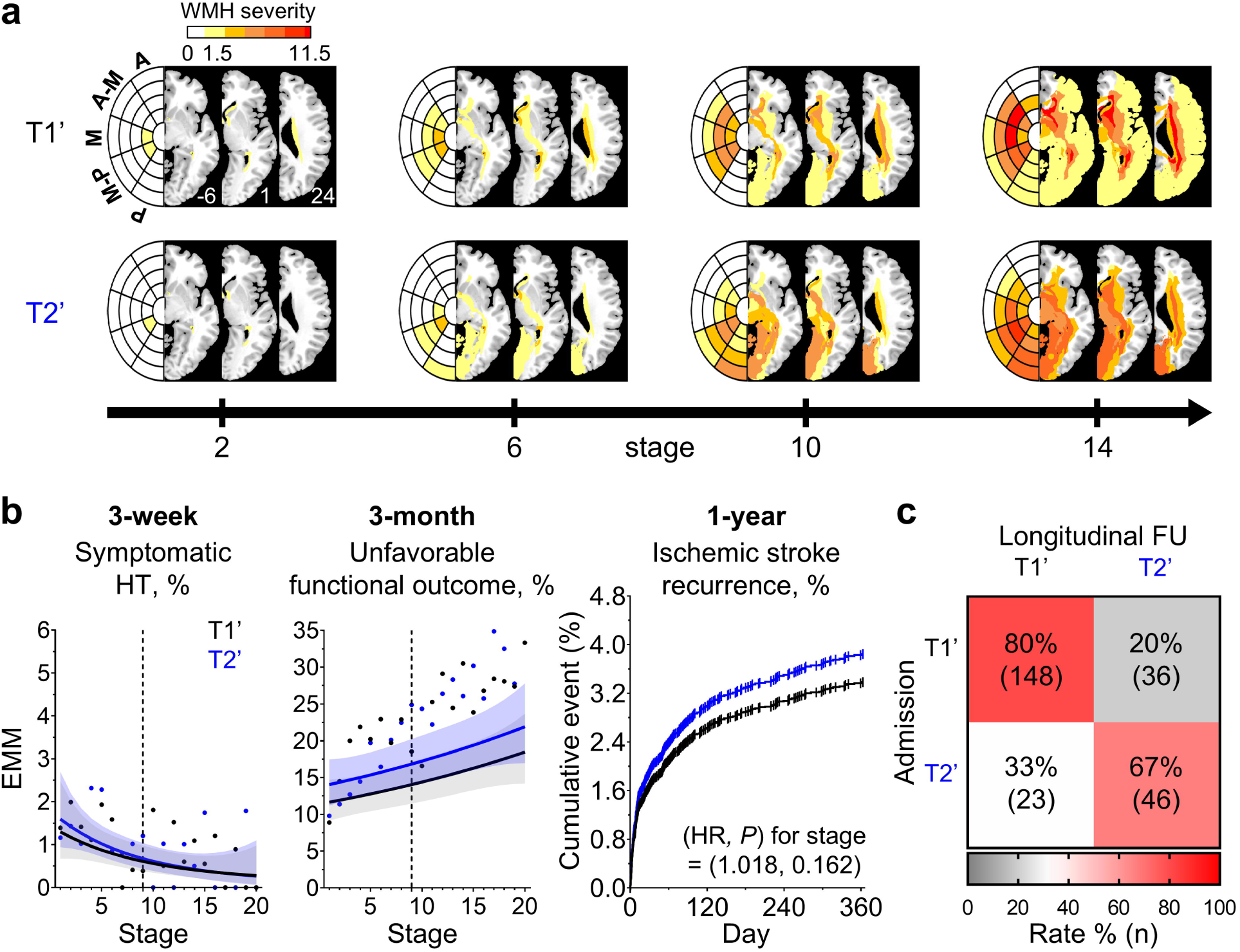
Cerebral arterial territory model for spatiotemporal patterns of white matter hyperintensity (WMH) progression in stroke patients. **a**, Spatiotemporal patterns of WMH progression across arterial territories (anterior cerebral artery [A], middle cerebral artery [M], posterior cerebral artery [P], and their border zones [A-M and M-P]). Bullseye presentations and median WMH severity maps are presented for each WMH subtype at WMH stages 2, 6, 10, and 14. The z-axis coordinates in the Montreal Neurological Institute space are displayed in the bottom right corner of the first three brain slices in the T1 group. **b**, Estimated marginal means (EMMs; a curve line with a shaded 95% confidence interval) of proportions of symptomatic hemorrhagic transformation (HT) leading early neurological deterioration within 3 weeks and unfavorable functional outcome at 3 months (modified Rankin Scale score > 3) by stage across subtypes (T1’ and T2’) are presented. Cumulative events of ischemic stroke recurrence within 1 year across subtypes are shown, as are hazard ratio (HR) and *P* for WMH stage. **c**, Longitudinal subtype stability rates by subtype at admission and follow-up (FU).

### Using propensity score matching for sensitivity analysis on WMH trajectories

To minimize potential modeling bias arising from population differences between stroke patients and low-risk controls, we applied propensity score matching and conducted SuStaIn modeling on the matched stroke patient cohort (Supplementary Fig. 1). The analysis included 7,311 stroke patients and 7,311 age- and sex-matched low-risk controls. A total of 412 (6%) patients were assigned as stage 0 and not classified to any subtype. The sensitivity analysis yielded results that were consistent with main analysis findings in terms of the three distinct spatiotemporal patterns of WMH progression (Supplementary Fig. 9a, b), baseline characteristics of the three subtypes (Supplementary Fig. 9c-g and Supplementary Table 11), and their post-stroke outcomes (Supplementary Fig. 10 and Supplementary Table 12). Unlike in the main results, T2 and T3 showed a higher incidence of nonvascular death than T1 (Supplementary Fig. 10c).

## Discussion

We identified distinct spatiotemporal trajectories of WMH progression by using statistical modeling with longitudinal inference^37^ based on a large cross-sectional multi-center dataset of consecutive stroke patients (*n* = 9,179). Consecutive enrollment and large sample size allowed us to reliably estimate WMH progression trajectories from cross-sectional data. Different WMH progression subtypes showed: i) distinct demographic and vascular risk factor profiles (delayed WMH onset, more women, and more frequent hypertension in fronto-parietal T1 vs. earlier WMH onset, more men, and more frequent AF in temporo-occipital T3); ii) distinct stroke etiology-related features (more frequent SVO in T1 vs. more frequent CE in T3); and iii) distinct patterns of stroke outcomes (more frequent early symptomatic hemorrhagic transformation and 3-month unfavorable functional outcome in T3 vs. more frequent 1-year ischemic stroke recurrence in T1). These data complemented or accorded with total or regional Fazekas scale-related findings. Longitudinal validation confirmed that the modeling closely aligned with observed WMH progression trajectories. Furthermore, the concordant WMH progression trajectories between high-risk controls and stroke patients, the divergence in their WMH stage distributions, and the consequent higher area under the curve in the ROC (compared to WMH volume) for distinguishing stroke patients from high-risk controls warrant further investigation to confirm whether WMH stage can better predict future stroke occurrence.

T1 (fronto-parietal WMH progression) was characterized by older age or late onset of severe WMH as well as higher proportions of female patients, hypertension, silent brain infarcts, and cerebral microbleeds. A previous study^50^ has also reported anterior-dominant distribution of WMH in older and hypertensive adults. Another study^33^ has shown that patients who are older, female, hypertensive, or have SVO tend to have more extensive WMH in the ACA territory than in MCA and PCA territories. T1 patients had the highest incidence of 1-year ischemic stroke recurrence, which might be partly attributed to their older age and higher prevalence of hypertension^51–53^. In addition, our study revealed that subtype differences in age distribution and hypertension prevalence were greater in early stages than in late stages. Differences in fronto-parietal WMH severity across subtypes were more pronounced in early stages. In late stages, T2 and T3 also showed WMH progression in fronto-parietal regions, likely due to age and hypertension. As a result, late-stage T2 and T3 patients might also be older and have higher rates of hypertension, similar to late-stage T1 patients.

T2, characterized by radial WMH progression across all lobes, was the predominant subtype, accounting for 46% of stroke patients in our study. Notably, vascular risk factors such as hypertension, AF, and coronary artery disease had the lowest prevalence in T2. This subtype had the highest prevalence of SVO. Obviously, both acute infarct volume and NIHSS score were lowest in T2 compared to the other subtypes.

T3 (temporo-occipital WMH progression) had the highest proportions of men and AF, along with the largest acute infarct volumes and the highest NIHSS scores. A previous study has found that temporal and posterior WMH is more frequently observed in patients with AF than in those without AF^54^. Moreover, CE strokes^55^ predominantly caused by AF are strongly associated with larger acute infarct volumes and elevated NIHSS scores^56,57^. Additionally,, earlier studies have shown that AF, larger acute infarct volume, and higher admission NIHSS score are associated with increased risk of symptomatic hemorrhagic transformation leading to early neurological deterioration^58,59^. These findings aligned with the relatively higher incidence of symptomatic hemorrhagic transformation and early neurological deterioration in T3 observed in our study, a connection which suggests T3 WMH might have potential clinical implications, particularly in situations, such as revascularization therapy, where hemorrhagic complications are a concern^60^. The incidence of unfavorable functional outcome at 3 months rose with advancing WMH stages across all subtypes but was highest in T3, likely due to larger acute infarct volume, higher NIHSS score, and more frequent hemorrhagic transformation.

The Fazekas scale, the most widely used WMH staging measure, can estimate overall WMH burden and predict stroke occurrence and prognosis^25,61,62^. Additionally, regional Fazekas scales for PVWMH and DWMH have been applied based on the hypothesis that PVWMH and DWMH have different vascular pathologies. However, it has been challenging to clearly differentiate regionally distinct pathophysiological mechanisms^5,63–65^.

Previous studies have shown that total Fazekas scale score is associated with the majority of risk factors related to both PVWMH and DWMH^5,63,64,66,67^, suggesting that PVWMH and DWMH likely represent a continuum of the same pathology. In a similar vein, there was a strong correlation between PVWMH and DWMH scores^5,63,64^, a finding the present study also corroborates (R^2^ = 0.50). We also showed that both total and PVWMH Fazekas scale scores were associated with age, female sex, hypertension, diabetes mellitus, AF, the presence of silent brain infarcts and cerebral microbleeds, stroke subtype (SVO and CE), acute infarct volume, and admission NIHSS score. The DWMH score was associated with age and hypertension—major risk factors for WMH—as well as the presence of silent brain infarcts and cerebral microbleeds. In our model, temporal information about WMH stages mirrored the relationships total and PVWMH Fazekas scale scores have with demographic and cerebrovascular risk factors, stroke etiology, and stroke severity. Additionally, our model provided the following spatial information with distinct WMH progression trajectories: i) older, hypertensive, and SVO patients were more likely to be T1 (fronto-parietal); ii) AF, coronary artery disease, and CE patients were more likely to be T3 (temporo-occipital); and iii) most clinical factors were less prevalent in T2 (radial).

In terms of stroke outcomes, total Fazekas scale score, but not PVWMH or DWMH scores, and WMH stage in our model were both associated with early neurological deterioration and its causes, including stroke progression and symptomatic hemorrhagic transformation. Moreover, unlike our WMH subtyping–staging model, total and regional Fazekas scale scores did not significantly predict poor functional outcome at 3 months. This further suggests that the new WMH subtyping–staging model aligns with and complements the conventional Fazekas scale in profiling stroke risk and outcomes.

Intriguingly, total and PVWMH Fazekas scale scores as well as WMH stage in our model all showed negative associations with the prevalence of CE etiology. Previous studies have reported that patients with larger WMH volumes are more likely to experience SVO strokes than those with other stroke subtypes, particularly CE^68,69^, which could partly explain the negative associations observed in the present study. We also found that only DWMH Fazekas scale score did not show a significant association with diabetes mellitus or AF, although previous studies have reported inconsistent associations between these factors and both PVWMH and DWMH^66,70–72^.

Our WMH subtyping–staging model offers a more refined spatiotemporal resolution for assessing WMH burden, encompassing 21 stages (0-20) for each of the three regional progression subtypes. This contrasts with the Fazekas scale, which has total scores ranging from 0-6, with 0-3 assigned to each of the two regional subtypes. Such increased granularity may enhance sensitivity in detecting small yet significant changes in WMH severity, thereby improving prognostic accuracy and contributing to personalized medicine. While the Fazekas scale is limited by fewer staging categories and qualitative assessment, it has been widely adopted due to its simplicity: anyone can easily assess it by visually inspecting images. Compared to the Fazekas scale, our model involves more complex steps, including the following processes: 1) delineating WMH based on brain ROIs, 2) constructing a statistical model, and 3) assigning individuals to specific subtypes and stages based on the model. These processes require a modest amount of computer processing, which currently limits user accessibility. However, this challenge could be addressed by developing an automated algorithm and implementing it with software. Automating the pipeline would allow users to fully benefit from the model without the current complexity. Our research team is actively developing this automated pipeline by using deep-learning algorithms. We plan to release the software before long, thus enabling users to easily analyze individual WMH subtype and stage directly from raw images. Through these efforts, we aim to maximize the strengths of our model and boost its applicability.

Our proposed WMH stage was associated with various demographics and vascular risk factors, including age, male sex, hypertension, diabetes mellitus, AF, coronary artery disease, and chronic SVD markers such as silent brain infarcts and microbleeds. These findings were largely consistent with previous studies using the Fazekas scales or WMH volume^66,68,69^. Interestingly, however, we found a positive association between male sex and WMH stage but a negative association between AF and WMH stage. The observed association between male sex and WMH stage might be attributed to an overcompensation for older female patients in the multivariate analysis. Regarding the inverse relationship between AF and WMH stage, as we previously discussed when reporting similar findings in elderly hypertensive patients^11^, AF could cause ischemic stroke (i.e., CE stroke) in the absence of other vascular risk factors that could contribute to elevated WMH stage^73^. Moreover, because our study found fewer CE strokes at higher WMH stages, and previous studies demonstrated that hemorrhagic transformation tended to occur more frequently in CE strokes (vs. strokes from other etiologies)^74^, it seems reasonable that the incidence of symptomatic hemorrhagic transformation decreased as WMH stages increased. Supporting this, the trend reversed and increased after we adjusted for stroke etiology (OR [95% CI]: 1.17 [1.12–1.21], *P* < 0.001).

We included only patients with first-ever stroke whose WMH were not attributable to clinically evident prior stroke, with FLAIR MRI scans acquired early (median [IQR] = 14 [6□38] hours) after symptom onset. Additionally, we have made great efforts to enroll patients consecutively to minimize selection bias. Despite potential inherent differences between stroke patients and high-risk controls, as well as racial differences between the two datasets used (i.e., the Korean MRI-based stroke database and the UK Biobank), we observed concordant spatiotemporal WMH progression trajectories in both groups. These findings might suggest that our subtyping–staging model may not only be applicable to stroke populations but could also seamlessly extend into a continuum within high-risk normal populations. In other words, our model could be broadly applicable as a potentially generalizable biomarker for cerebrovascular brain health. Future studies should confirm whether our current findings in a first-ever stroke patient population can be extrapolated to high-risk, stroke-naïve populations.

In our study, the mean [95% CI] WMH volume (mL) in the UK Biobank controls was 4.20 [4.14□4.26], which closely aligns with findings from a meta-analysis of 17 studies involving 9,716 healthy subjects^75^. In both high-risk and low-risk controls, WMH volumes showed right-skewed distribution, and this result is consistent with the typical right-skewed distribution of WMH volumes observed in population-based cohorts^76,77^. Stroke patients from our Korean MRI-based stroke database had less right-skewed and significantly higher WMH volumes (Median [IQR]: 8.90 [4.39□18.17]) compared to UK Biobank controls (2.45 [1.61□4.37]), in accordance with previous studies on stroke patients^23,33,68,69^. Since the UK Biobank represents a general (low- and high-risk) control population and the Korean MRI-based stroke database represents a stroke population, the significantly different stage distributions between these groups have notable implications for stroke risk assessment, particularly in high-risk individuals.

High-risk controls were predominantly concentrated in the early stages and had significantly more left-skewed distribution than stroke patients. This finding suggests the model can estimate subclinical WMH burden that is a precursor to symptomatic stroke. Previous longitudinal studies have shown that larger baseline WMH volumes were associated with elevated stroke risk, with hazard ratios ranging from 1.4 to 1.79 per log-unit increase in WMH volume^78,79^. In our ROC analysis, WMH stage demonstrated significantly greater discriminative power than WMH volume for distinguishing stroke patients from high-risk controls. These results indicate that the WMH stage derived from our WMH subtyping– staging model provides a more precise and reliable tool for predicting symptomatic stroke risk compared to traditional WMH volume-based approaches.

The WMH subtypes and stages identified based on functional lobes effectively predicted post-stroke outcomes. However, the alternative WMH progression model based on arterial territories did not effectively predict symptomatic hemorrhagic transformation, 3-month unfavorable functional outcomes, or 1-year ischemic stroke recurrence. Moreover, when reassessed at an FU time point in the same patients, the WMH subtypes were more likely to show changes and are thus less reliable predictors of WMH progression trajectories. Because WMH is closely linked to vascular risk factors, arterial territories would be expected to be a better basis for modeling (compared to functional lobes of the brain^8,33,80^), yet our findings suggest that pathological processes of WMH (a marker of SVD) might be less affected by mechanisms related to large artery disease.

This study has some limitations. We used data from low-risk controls in the UK Biobank to normalize WMH volumes into z-scores (WMH severity) for each ROI in stroke patients from the Korean MRI-based stroke database. Since the UK Biobank dataset primarily consisted of Caucasian individuals with only a small proportion of Asians, we made efforts to minimize bias due to differences in the datasets’ racial composition. We constructed a low-risk control dataset from the UK Biobank, excluding individuals with hypertension, diabetes mellitus, hyperlipidemia, current smoking, AF, or coronary artery disease, to mitigate racial differences in the effects vascular risk factors exert on WMH progression. Additionally, we conducted a sensitivity analysis using propensity score matching for demographics, such as age and sex, between datasets. Repeating our main analyses with these matched datasets produced nearly identical results. Nevertheless, a previous study has shown that, even after controlling for demographics and risk factors, a Chinese population had a higher prevalence of severe WMH than a white population^81^. Thus, caution should be exercised when applying these findings to other racial groups. Further validation with a large racially matched dataset should be performed in future studies.

In conclusion, we developed and validated a new WMH subtyping–staging model by identifying distinct spatiotemporal trajectories of WMH progression, and we demonstrated that this model can not only reliably reflect demographics and vascular risk factors but also predict stroke outcomes across various WMH subtypes and stages. Moreover, the aligned WMH progression trajectories observed in high-risk controls and stroke patients, combined with their distinct stage distributions, suggest this model could potentially predict stroke risk in the general population.

## Methods

All analyses were conducted using MATLAB R2021b (MathWorks, Natick, MA, USA), unless otherwise noted.

### Dataset

The flow diagram of the study data is presented in Supplementary Fig. 1. The main data on stroke patients (*n* = 9,179) were obtained from the Korean MRI-based stroke database^11,40–42^, a subproject of Clinical Research Collaboration for Stroke-Korea (CRCS-K, a nationwide stroke registry; http://crcs-k.strokedb.or.kr/eng/). The database consecutively enrolled patients (*n* = 13,186) with acute stroke who were admitted to 11 participating centers within 7 days of symptom onset between May 2011 and February 2014. Data from one center (*n* = 748) were separated and assigned to validation data. Patients (*n* = 4,007 in the main data, *n* = 191 in the validation data) were excluded based on the following criteria: non-ischemic stroke, previous stroke history, poor-quality FLAIR MRI, and no white matter hyperintensity in FLAIR MRI. The validation data were further refined by excluding 280 patients without FU FLAIR MRI and 13 patients with poor-quality FU FLAIR MRI, resulting in a total of 264 patients. This study was approved by the institutional review boards of all participating centers (DUIH2010-01-083-020).

We collected demographic and clinical data, including medication history, laboratory results, and vascular risk factors, by using a standardized protocol^11,40^. Acute ischemic stroke subtypes were determined by consensus among experienced vascular neurologists at each center, all using a validated MRI-based algorithm^82^ based on the Trial of Org 10172 in Acute Stroke Treatment (TOAST) classification^83^. The mRS score^84^ before and 3 months after stroke (FU loss in *n* = 229) and NIHSS score^85^ at admission were collected prospectively. Neurological worsening and new neurological symptoms within 3 weeks of stroke onset (defined as early neurological deterioration) were assessed using previously published criteria^40,42,86,87^ (FU loss in *n* = 4). Data quality was ensured through regular audits, including monthly monitoring and on-site inspections to review medical records, conducted by an outcome adjudication committee. All patients underwent standard clinical evaluation, treatment, and rehabilitation in accordance with current guidelines for stroke, as published in CRCS-K annual report: http://crcs-k.strokedb.or.kr/eng/reporting/report.asp. Stroke recurrence and mortality within 1 year were prospectively collected using a predefined protocol^88^ during hospitalization, or after discharge through routine clinic visits or telephone interviews (FU loss in *n* = 93). Vascular death was defined as any death that occurred during the qualifying stroke admission period, including fatal stroke recurrence, fatal myocardial infarction, fatal congestive heart failure, and any sudden death without an identifiable nonvascular cause after discharge. Nonvascular death was defined as any death not caused by vascular causes.

From the UK Biobank database^89^, we initially included individuals (*n* = 37,070) with both (baseline) T1-weighted and FLAIR MRI scans in this study. For control data, individuals with neurological diseases or without white matter hyperintensity in FLAIR MRI were excluded (*n* = 861). Only individuals with no vascular risk factors, including hypertension, diabetes mellitus, hyperlipidemia, current smoking, AF, and coronary artery disease, were included (*n* = 13,811) to construct a “low-risk control” population in relation to stroke. These low-risk control data were used to estimate baseline distributions of WMH volumes in each ROI, and these estimated distributions were then used to calculate WMH severity as z-scores. Binarized diagnoses were determined by assessing all relevant data fields (ex: ICD10, self-report, medications). Diagnoses included hypertension (data field number: 41270, 20002, 6150), diabetes (41270, 20002, 6153, 6177), hyperlipidemia (41270, 20002), smoking status (20116), AF (41270, 41272, 20002, 20004, 12653), and coronary artery disease (41270, 41272, 20002, 20004, 6150).

### MRI acquisition and processing

Multicenter MRI protocols for stroke patient data were as follows. Diffusion-weighted MRI was acquired with b-values of 0 and 1,000 s/mm^2^, 2400–10400 ms repetition time, 47–111 ms echo time, 0.6–2×0.6–2×3–6 mm^3^ voxel size, 3–7.5 mm interslice gap, and 200– 260×200–260 mm FOV. FLAIR MRI was acquired with 6000–13000 ms repetition time, 76– 169 ms echo time, 0.3–0.9×0.3–0.9×5–6 mm^3^ voxel size, 3–7.5 mm interslice gap, and 160– 250×200–250 mm FOV. T2-weigthed MRI was acquired with 1500–8800 ms repetition time, 45–126 ms echo time, 0.2–0.9×0.2–0.9×3–6 mm^3^ voxel size, 3–7.5 mm interslice gap, and 160–250×200–250 mm FOV. Gradient-echo MRI was acquired with 350–1230 ms repetition time, 14–30 ms echo time, 0.4–0.9×0.4–0.9×3–6 mm^3^ voxel size, 4–7.5 mm interslice gap, and 160–250×200–250 mm FOV. As previously described^11,40^, stroke-related lesions on MR images were semi-automatically segmented and registered onto a standard brain template^90^ in Montreal Neurological Institute (MNI) space under slice-by-slice supervision by an experienced vascular neurologist (author W.-S.R.). The lesion quantification method demonstrated excellent inter- and intra-observer reliability^11^.

UK Biobank structural imaging included 3D MPRAGE T1-weighted MRI acquisition with 1 mm isotropic resolution (repetition time = 2000 ms, echo time = 2 ms, slice thickness = 1 mm, FOV = 192×256×256) and 3D SPACE FLAIR MRI acquisition with 1.05×1×1 mm resolution (repetition time = 5000 ms, echo time = 395 ms, slice thickness = 1.05 mm, FOV = 192×256×256). T1-weighted MRI was preprocessed using FreeSurfer^91^ v7.1 and the bias-corrected image was linearly registered to the FLAIR using six degrees of freedom. Both T1-weighted and FLAIR images were used as inputs for WMH segmentation following the method described by Park et al., 2021^92^. This deep-learning based algorithm, an ensemble U-Net with multi-scale highlighting foregrounds, demonstrated exceptional performance in detecting WMH and placed first among 57 teams in the WMH segmentation challenge (https://wmh.isi.uu.nl/#_Results) ^93^. The segmented WMH lesion in the native space was then registered to the standard brain template in MNI space by using a non-linear registration method (FMRIB Software Library [FSL], FNIRT^94^).

### Brain parcellation

For main analyses, we used a bullseye parcellation method^31,95^ to define white matter ROIs for WMH. We applied the bullseye pipeline (https://github.com/gsanroma/bullseye_pipeline) to the standard brain template in MNI space, segmenting the brain into 20 ROIs (Supplementary Fig. 2). Each functional lobe, including frontal, basal ganglia, temporal, parietal, and occipital lobes, was divided into four layers based on the relative distance to lateral ventricles and cortical surfaces: layer 1 (closest to the ventricles) ranged from 0 to 0.25, layer 2 ranged from 0.25 to 0.5, layer 3 ranged from 0.5 to 0.75, and layer 4 (closest to the cortical surfaces) ranged from 0.75 to 1. We chose bullseye parcellation for its excellent spatial resolution across axial, sagittal, and coronal planes of brain, plus its capacity to incorporate conventional parcellation based on distance from ventricles. Additionally, we generated another bullseye parcellation using cerebral arterial territories instead of functional lobes (Supplementary Fig. 8). We have previously specified cerebral arterial territories of major arteries (ACA, MCA, and PCA) and their border zones (ACA-MCA and MCA-PCA) in MNI space^41^. We again divided each arterial territory into four layers, resulting in a total of 20 ROIs.

### SuStaIn modeling

SuStaIn modeling is event-based modeling^96,97^ that describes disease progression as a sequence of discrete events, wherein the total number of stages equals the number of events. Each event is defined as the incident at which a disease-related biomarker reaches a predefined abnormal level (cutoff). For a detailed description of the mathematical framework, see previously published work by Young et al.^94^. In the present study, to identify imaging biomarkers (i.e., subtyping and staging) for spatiotemporal WMH progression we estimated subtype-specific WMH severity evolution by using cross-sectional datasets, in which each individual contributed a single severity value derived from 20 prespecified ROIs. Thus, the number of total WMH progression stages was set to 20. We evaluated abnormal levels by calculating ROI-wise WMH severity in stroke patients (from the Korean MRI-based stroke database) relative to a low-risk control subpopulation (from the UK Biobank). First, we calculated ROI-wise WMH volumes in both low-risk controls and stroke patients by counting lesioned voxels. Poisson regression was performed on WMH volumes in each ROI for the low-risk controls, with age as an independent variable. The effect of age was then regressed from raw WMH volumes in each ROI to obtain age-adjusted WMH volumes for low-risk controls. The mean and standard deviation of these age-adjusted WMH volumes in low-risk controls were then used to normalize raw WMH volumes in stroke patients, thereby yielding ROI-specific WMH severities (z-scores). Regressions and normalizations were performed separately for men and women.

We applied SuStaIn modeling to the stroke patient dataset, with a WMH severity (z-scores) cutoff set to 1.5 and the maximum WMH severity threshold set to the median of 95th percentile values derived from ROI-wise WMH severities in stroke patients (8.5 for the main model). Model fitting and uncertainty estimation methods were adopted from previous work by Young et al.^94^. The expectation-maximization algorithm for model fitting was run with 25 different start points for random cluster assignments. Model uncertainty was then estimated through 100,000 Markov chain Monte Carlo iterations. The maximum number of subtypes was set to five. The optimal number of subtypes was decided by 10-fold CV. Out-of-sample model fits were evaluated by calculating the log-likelihood and CVIC^37,38^ across each N-subtype model. Progression pattern similarities within the same subtype between CV folds were assessed using the Kendall rank correlation coefficient for estimated event sequences. Similarly, progression pattern similarities across different subtypes were assessed using the Kendall rank correlation coefficient within each CV fold, and the maximum similarity for all subtype pairs was identified within each fold. We determined the optimal number of subtypes by considering all CV evaluation metrics. Then, individual subtypes were estimated by integrating likelihood across all stages and selecting the subtype with the highest likelihood. Each individual’s stage was then assigned based on the stage with the highest likelihood for the selected subtype. All likelihoods were integrated across all Markov chain Monte Carlo samples. Probability of ML subtype was calculated as the likelihood of the assigned subtype divided by the sum of all subtype likelihoods for each individual.

### Prediction of longitudinal change in WMH stage

We applied linear multivariable regression models with leave-one-out CV to the validation data in order to predict the change in WMH stage difference from admission to FU. Models included FU duration (in years); baseline characteristics such as age, sex, hypertension, diabetes mellitus, hyperlipidemia, current smoking, AF, coronary artery disease, and revascularization therapy; plus WMH subtype and WMH stage at admission. After calculating predicted changes in WMH stage, we performed a linear regression analysis comparing observed WMH stage changes to predicted changes.

### Subtype characterization of demographics, vascular risk factors, stroke etiology, stroke severity, and post-stroke outcomes

Subtype differences in demographics, vascular risk factors, and all stroke outcomes were evaluated. We used mixed-effects regression models—linear regression for continuous variables and logistic regression for categorical variables—to analyze all clinical assessments, except 1-year stroke recurrence and mortality, as dependent variables (outcomes). The mixed-effects regression model for each clinical assessment included age (except in the case of age), sex (except in the case of sex), subtype, stage, and subtype–stage interaction as fixed effects, with imaging center as a random effect. Inclusion of subtype-stage interaction was determined by comparing restricted ML (for continuous variables) and Akaike information criterion (for categorical variables) between models with and without the interaction term. Even if the model with the interaction term provided a better fit, the model without the interaction term was finally selected if the *P* for the interaction exceeded 0.1. When subtype– stage interaction was included in the model, subgroup analyses were extended by dividing patients into stage ≤ 9 (median) or stage > 9 to further evaluate subtype differences in clinical assessments along stage. Onset to MRI time was included as a fixed factor for acute infarct volume analysis. Revascularization therapy was included as a fixed factor for post-stroke outcomes such as early neurological deterioration within 3 weeks, early neurological deterioration causes, and unfavorable functional outcome at 3 months. For 3-month unfavorable functional outcome, only patients with pre-stroke mRS scores < 2 were included, with pre-stroke mRS score added as a covariate. EMMs across stages for each dependent variable were calculated by marginal effects at average values of covariates^98^. Acute infarct volume and NIHSS scores (as dependent variables) were log-transformed to account for skewed distributions that caused model residuals to deviate from normality. EMMs and Cis were back-transformed to their original scale.

We performed cox proportional hazards regression models for 1-year stroke recurrence and mortality. These models included WMH subtype and stage with covariates such as age, sex, and revascularization therapy. Inclusion of subtype–stage interaction was determined by comparing Akaike information criterion between models, with the interaction term excluded if its *P* exceeded 0.1.

### Fazekas scale assessment

We assessed Fazekas scale in 9,179 stroke patients from the main dataset. Five experienced vascular neurologists visually rated PVWMH and DWMH on a 0–3 scale, each for approximately 2,000 patients. All raters followed the same instructions, based on the report by Fazekas et al. in 1987^30^. To evaluate inter-rater reliability, all raters assessed the same 100 patients. Intraclass correlation coefficient values and 95% CIs for total, PVWMH, and DWMH Fazekas scales were 0.97 [0.96–0.98], 0.95 [0.93–0.96], and 0.95 [0.93–0.96], respectively.

### Discrimination between high-risk controls and stroke patients by WMH stage vs. volume

We compared the distributions of WMH stage and volume between high-risk controls and stroke patients by using Mann–Whitney U-tests. Next, we generated ROC curves for each measure (WMH stage and volume) to assess their capability to discriminate between the two groups, using the “pROC” package in R software (version 4.3.1). Finally, we compared the areas under the ROC curves by using DeLong test^99^.

### Analysis of trajectory deviations in spatiotemporal WMH progression between stroke patients and high-risk controls

We performed PCA, an unsupervised dimensionality reduction technique^47–49^, to construct a representative trajectory space for WMH maps of the stroke group (Fig. 1a) by identifying PCs that capture the most variance across different WMH subtypes and stages, while minimizing trajectory-unrelated variances. The maps, which depict median WMH severity for each of the 20 ROIs at 20 stages and three subtypes, could be expressed as a set of 60 (20 stages × 3 subtypes) coordinates in a 20-dimensional space, where each dimension corresponds to a specific ROI. We selected the first three PCs in descending order of their variance explained, which collectively accounted for 95% of the total variance, to construct the representative trajectory space. Using the three PCs derived from the stroke group data, we mapped the 60 coordinates for the stroke group and the high-risk control group into a shared representative three-dimensional trajectory space. Finally, each PC was z-normalized using the mean and standard deviation derived from the combined transformed data of both stroke patients and high-risk controls. To estimate trajectory deviations in WMH spatiotemporal progression in the high-risk control group compared to the stroke group, we calculated the Euclidean distances between the three-dimensional coordinates of individual high-risk controls and the corresponding median coordinates of the stroke group for each stage and subtype, thereby generating 60 (20 stages × 3 subtypes) median values with their 95^th^ percentiles. Additionally, we computed within-population 95^th^ percentiles for the stroke group. For each subtype and stage, outliers among high-risk controls were identified using the 95^th^ percentile of Euclidean distances observed in the stroke group.

### Cerebral arterial territory model for WMH progression

SuStaIn modeling was performed for stroke patients by using cerebral arterial territory parcellation (Supplementary Fig. 8), with WMH severity cutoff set to 1.5 and maximum WMH severity set to 11.5 (the median of the 95^th^ percentiles of ROI-wise WMH severities in stroke patients). We repeated the same procedure to evaluate the cerebral arterial territory model as the main model.

### Sensitivity analysis

We used propensity score matching to extract age- and sex-matched low-risk controls and stroke patients (Supplementary Fig. 1). Propensity scores were calculated using logistic regression based on age and sex, followed by 1:1 matching with a caliper width of 0.2. After matching, 7,311 stroke patients and 7,311 low-risk controls were included for sensitivity analysis, which was performed using MatchIt^100^ and cobalt^101^ packages in R software version 4.3.1. We repeated the same analyses as for the main model.

## Data availability

Anonymized data from the Korean MRI-based stroke database used in this study can be made available upon reasonable request, subject to approval by the CRCS-K steering committee.

## Code availability

All preprocessing, statistical analysis, and visualization codes are openly available on GitHub (https://github.com/JinyongChung/WMH_progression_modeling). For SuStaIn modeling, we utilized source codes developed by Young et al.^94^, which are available at https://github.com/ucl-mig/SuStaInMatlab. Our repository includes the constructed WMH progression model and the codes for determining individual subtypes and stages based on WMH severity. Researchers can apply our methodology to their own data by calculating WMH severity by using the specified ROIs employed in this study. The ROI templates, which define the specified ROIs in the MNI space, are also included in the repository.

## Acknowledgements

This research was conducted using data from the UK Biobank (www.ukbiobank.ac.uk). This research was approved by the UK Biobank (application number: 11559). The study authors appreciate contributions from all CRCS-K members. This study was supported by grants from the National Priority Research Center Program (NRF-2021R1A6A1A03038865) and Bioimaging Data Curation Center Program (NRF-2022M3H9A2096198) of the National Research Foundation funded by the Korean government.

## Author contributions

D.K. supervised this study and acquired the funding. J.C., W.R., and D.K. designed the study and planned the methodology. D.K. and H.B. developed, and W.R., B.J.K., K.H., S.J., J.K., K.L., T.H.P., S.P., J.P., K.K., H.P., Y.C., B.L., K.Y., M.S.O., S.J.L., J.G.K., J.C., D.K., J.L., M.H., M.S.P., and K.C. contributed to, the Korean MRI-based stroke database. J.C., G.P., E.H., and D.K. performed data analysis. J.C., G.P., W.R., D.S., H.K., D.G., H.O., H.K., and D.K. performed data interpretation and visualization. J.C., G.P., E.H., and D.K. wrote the original draft. All authors contributed to the manuscript revision and review.

## Additional information

Supplemental information.

## References

1 de Leeuw, F. E. et al. Prevalence of cerebral white matter lesions in elderly people: a population based magnetic resonance imaging study. The Rotterdam Scan Study. J Neurol Neurosurg Psychiatry 70, 9–14 (2001). 10.1136/jnnp.70.1.9

2 Debette, S. & Markus, H. S. The clinical importance of white matter hyperintensities on brain magnetic resonance imaging: systematic review and meta-analysis. BMJ 341, c3666 (2010). 10.1136/bmj.c3666

3 Habes, M. et al. White matter hyperintensities and imaging patterns of brain ageing in the general population. Brain 139, 1164–1179 (2016). 10.1093/brain/aww008

4 Hopkins, R. O. et al. Prevalence of white matter hyperintensities in a young healthy population. J Neuroimaging 16, 243–251 (2006). 10.1111/j.1552-6569.2006.00047.x

5 Kim, K. W., MacFall, J. R. & Payne, M. E. Classification of white matter lesions on magnetic resonance imaging in elderly persons. Biol Psychiatry 64, 273–280 (2008). 10.1016/j.biopsych.2008.03.024

6 Launer, L. J. et al. Regional variability in the prevalence of cerebral white matter lesions: an MRI study in 9 European countries (CASCADE). Neuroepidemiology 26, 23–29 (2006). 10.1159/000089233

7 Prins, N. D. & Scheltens, P. White matter hyperintensities, cognitive impairment and dementia: an update. Nat Rev Neurol 11, 157–165 (2015). 10.1038/nrneurol.2015.10

8 Wardlaw, J. M., Valdes Hernandez, M. C. & Munoz-Maniega, S. What are white matter hyperintensities made of? Relevance to vascular cognitive impairment. J Am Heart Assoc 4, 001140 (2015). 10.1161/JAHA.114.001140

9 Zhuang, F. J., Chen, Y., He, W. B. & Cai, Z. Y. Prevalence of white matter hyperintensities increases with age. Neural Regen Res 13, 2141–2146 (2018). 10.4103/1673-5374.241465

10 Garnier-Crussard, A. et al. White matter hyperintensities across the adult lifespan: relation to age, Abeta load, and cognition. Alzheimers Res Ther 12, 127 (2020). 10.1186/s13195-020-00669-4

11 Ryu, W. S. et al. Grading and interpretation of white matter hyperintensities using statistical maps. Stroke 45, 3567–3575 (2014). 10.1161/STROKEAHA.114.006662

12 Ay, H. et al. Severity of leukoaraiosis and susceptibility to infarct growth in acute stroke. Stroke 39, 1409–1413 (2008). 10.1161/STROKEAHA.107.501932

13 Baik, M. et al. Differential impact of white matter hyperintensities on long-term outcomes in ischemic stroke patients with large artery atherosclerosis. PLoS One 12, e0189611 (2017). 10.1371/journal.pone.0189611

14 Bonkhoff, A. K. et al. Association of Stroke Lesion Pattern and White Matter Hyperintensity Burden With Stroke Severity and Outcome. Neurology 99, e1364–e1379 (2022). 10.1212/WNL.0000000000200926

15 Etherton, M. R. et al. White Matter Integrity and Early Outcomes After Acute Ischemic Stroke. Transl Stroke Res 10, 630–638 (2019). 10.1007/s12975-019-0689-4

16 Fu, J. H. et al. Extent of white matter lesions is related to acute subcortical infarcts and predicts further stroke risk in patients with first ever ischaemic stroke. J Neurol Neurosurg Psychiatry 76, 793–796 (2005). 10.1136/jnnp.2003.032771

17 Kang, H. J. et al. White matter hyperintensities and functional outcomes at 2 weeks and 1 year after stroke. Cerebrovasc Dis 35, 138–145 (2013). 10.1159/000346604

18 Kerber, K. A., Whitman, G. T., Brown, D. L. & Baloh, R. W. Increased risk of death in community-dwelling older people with white matter hyperintensities on MRI. J Neurol Sci 250, 33–38 (2006). 10.1016/j.jns.2006.06.022

19 Kissela, B. et al. Clinical prediction of functional outcome after ischemic stroke: the surprising importance of periventricular white matter disease and race. Stroke 40, 530–536 (2009). 10.1161/STROKEAHA.108.521906

20 Kumral, E. et al. Association of leukoaraiosis with stroke recurrence within 5 years after initial stroke. J Stroke Cerebrovasc Dis 24, 573–582 (2015). 10.1016/j.jstrokecerebrovasdis.2014.10.002

21 Ryu, W. S. et al. White matter hyperintensity load on stroke recurrence and mortality at 1 year after ischemic stroke. Neurology 93, e578–e589 (2019). 10.1212/WNL.0000000000007896

22 Botz, J., Lohner, V. & Schirmer, M. D. Spatial patterns of white matter hyperintensities: a systematic review. Front Aging Neurosci 15, 1165324 (2023). 10.3389/fnagi.2023.1165324

23 Wen, W. & Sachdev, P. The topography of white matter hyperintensities on brain MRI in healthy 60- to 64-year-old individuals. Neuroimage 22, 144–154 (2004). 10.1016/j.neuroimage.2003.12.027

24 Gouw, A. A. et al. Heterogeneity of small vessel disease: a systematic review of MRI and histopathology correlations. J Neurol Neurosurg Psychiatry 82, 126–135 (2011). 10.1136/jnnp.2009.204685

25 Gouw, A. A. et al. Heterogeneity of white matter hyperintensities in Alzheimer’s disease: post-mortem quantitative MRI and neuropathology. Brain 131, 3286–3298 (2008). 10.1093/brain/awn265

26 Lahna, D. et al. Venous Collagenosis as Pathogenesis of White Matter Hyperintensity. Ann Neurol 92, 992–1000 (2022). 10.1002/ana.26487

27 Li, Y. et al. The relationship between blood-brain barrier permeability and enlarged perivascular spaces: a cross-sectional study. Clin Interv Aging 14, 871–878 (2019). 10.2147/CIA.S204269

28 Walsh, J. et al. Microglial activation and blood-brain barrier permeability in cerebral small vessel disease. Brain 144, 1361–1371 (2021). 10.1093/brain/awab003

29 Zhang, W. et al. Glymphatic clearance function in patients with cerebral small vessel disease. Neuroimage 238, 118257 (2021). 10.1016/j.neuroimage.2021.118257

30 Fazekas, F., Chawluk, J. B., Alavi, A., Hurtig, H. I. & Zimmerman, R. A. MR signal abnormalities at 1.5 T in Alzheimer’s dementia and normal aging. AJR Am J Roentgenol 149, 351–356 (1987). 10.2214/ajr.149.2.351

31 Sudre, C. H. et al. Bullseye’s representation of cerebral white matter hyperintensities. J Neuroradiol 45, 114–122 (2018). 10.1016/j.neurad.2017.10.001

32 Phuah, C. L. et al. Association of Data-Driven White Matter Hyperintensity Spatial Signatures With Distinct Cerebral Small Vessel Disease Etiologies. Neurology 99, e2535–e2547 (2022). 10.1212/WNL.0000000000201186

33 Schirmer, M. D. et al. Spatial Signature of White Matter Hyperintensities in Stroke Patients. Front Neurol 10, 208 (2019). 10.3389/fneur.2019.00208

34 Cai, M. et al. Determinants and Temporal Dynamics of Cerebral Small Vessel Disease: 14-Year Follow-Up. Stroke 53, 2789–2798 (2022). 10.1161/STROKEAHA.121.038099

35 Jochems, A. C. C. et al. Longitudinal Changes of White Matter Hyperintensities in Sporadic Small Vessel Disease: A Systematic Review and Meta-analysis. Neurology 99, e2454–e2463 (2022). 10.1212/WNL.0000000000201205

36 Ramirez, J., McNeely, A. A., Berezuk, C., Gao, F. & Black, S. E. Dynamic Progression of White Matter Hyperintensities in Alzheimer’s Disease and Normal Aging: Results from the Sunnybrook Dementia Study. Front Aging Neurosci 8, 62 (2016). 10.3389/fnagi.2016.00062

37 Young, A. L. et al. Uncovering the heterogeneity and temporal complexity of neurodegenerative diseases with Subtype and Stage Inference. Nat Commun 9, 4273 (2018). 10.1038/s41467-018-05892-0

38 Vogel, J. W. et al. Four distinct trajectories of tau deposition identified in Alzheimer’s disease. Nat Med 27, 871–881 (2021). 10.1038/s41591-021-01309-6

39 Eshaghi, A. et al. Identifying multiple sclerosis subtypes using unsupervised machine learning and MRI data. Nat Commun 12, 2078 (2021). 10.1038/s41467-021-22265-2

40 Ryu, W. S. et al. Stroke outcomes are worse with larger leukoaraiosis volumes. Brain 140, 158–170 (2017). 10.1093/brain/aww259

41 Kim, D. E. et al. Mapping the Supratentorial Cerebral Arterial Territories Using 1160 Large Artery Infarcts. JAMA Neurol 76, 72–80 (2019). 10.1001/jamaneurol.2018.2808

42 Ryu, W. S. et al. Biological Mechanism of Sex Difference in Stroke Manifestation and Outcomes. Neurology 100, e2490–e2503 (2023). 10.1212/WNL.0000000000207346

43 Wardlaw, J. M., Smith, C. & Dichgans, M. Small vessel disease: mechanisms and clinical implications. Lancet Neurol 18, 684–696 (2019). 10.1016/S1474-4422(19)30079-1

44 Vermeer, S. E., Longstreth, W. T., Jr. & Koudstaal, P. J. Silent brain infarcts: a systematic review. Lancet Neurol 6, 611–619 (2007). 10.1016/S1474-4422(07)70170-9

45 Dufouil, C. et al. Longitudinal study of blood pressure and white matter hyperintensities: the EVA MRI Cohort. Neurology 56, 921–926 (2001). 10.1212/wnl.56.7.921

46 Scharf, E. L. et al. Cardiometabolic Health and Longitudinal Progression of White Matter Hyperintensity: The Mayo Clinic Study of Aging. Stroke 50, 3037–3044 (2019). 10.1161/STROKEAHA.119.025822

47 Ringner, M. What is principal component analysis? Nat Biotechnol 26, 303–304 (2008). 10.1038/nbt0308-303

48 Park, H., Yang, J. J., Seo, J., Lee, J. M. & Adni. Dimensionality reduced cortical features and their use in predicting longitudinal changes in Alzheimer’s disease. Neurosci Lett 550, 17–22 (2013). 10.1016/j.neulet.2013.06.042

49 Jolliffe, I. T. & Cadima, J. Principal component analysis: a review and recent developments. Philos Trans A Math Phys Eng Sci 374, 20150202 (2016). 10.1098/rsta.2015.0202

50 Wang, J. et al. Impact of different white matter hyperintensities patterns on cognition: A cross-sectional and longitudinal study. Neuroimage Clin 34, 102978 (2022). 10.1016/j.nicl.2022.102978

51 Chung, J. Y., Lee, B. N., Kim, Y. S., Shin, B. S. & Kang, H. G. Sex differences and risk factors in recurrent ischemic stroke. Front Neurol 14, 1028431 (2023). 10.3389/fneur.2023.1028431

52 Xu, J. et al. Trends and Risk Factors Associated With Stroke Recurrence in China, 2007-2018. JAMA Netw Open 5, e2216341 (2022). 10.1001/jamanetworkopen.2022.16341

53 Zheng, S. & Yao, B. Impact of risk factors for recurrence after the first ischemic stroke in adults: A systematic review and meta-analysis. J Clin Neurosci 60, 24–30 (2019). 10.1016/j.jocn.2018.10.026

54 Wiggins, M. E. et al. Pilot Investigation: Older Adults With Atrial Fibrillation Demonstrate Greater Brain Leukoaraiosis in Infracortical and Deep Regions Relative to Non-Atrial Fibrillation Peers. Front Aging Neurosci 12, 271 (2020). 10.3389/fnagi.2020.00271

55 Pistoia, F. et al. The Epidemiology of Atrial Fibrillation and Stroke. Cardiol Clin 34, 255–268 (2016). 10.1016/j.ccl.2015.12.002

56 Adams, H. P., Jr., et al. Baseline NIH Stroke Scale score strongly predicts outcome after stroke: A report of the Trial of Org 10172 in Acute Stroke Treatment (TOAST). Neurology 53, 126–131 (1999). 10.1212/wnl.53.1.126

57 Daneshvari, N. O. & Johansen, M. C. Associations between cerebral magnetic resonance imaging infarct volume and acute ischemic stroke etiology. PLoS One 16, e0256458 (2021). 10.1371/journal.pone.0256458

58 Fiorelli, M. et al. Hemorrhagic transformation within 36 hours of a cerebral infarct: relationships with early clinical deterioration and 3-month outcome in the European Cooperative Acute Stroke Study I (ECASS I) cohort. Stroke 30, 2280–2284 (1999). 10.1161/01.str.30.11.2280

59 Honig, A. et al. Hemorrhagic Transformation in Acute Ischemic Stroke: A Quantitative Systematic Review. J Clin Med 11 (2022). 10.3390/jcm11051162

60 Khatri, P., Wechsler, L. R. & Broderick, J. P. Intracranial hemorrhage associated with revascularization therapies. Stroke 38, 431–440 (2007). 10.1161/01.STR.0000254524.23708.c9

61 Georgakis, M. K., Duering, M., Wardlaw, J. M. & Dichgans, M. WMH and long-term outcomes in ischemic stroke: A systematic review and meta-analysis. Neurology 92, e1298–e1308 (2019). 10.1212/WNL.0000000000007142

62 Liou, L. M. et al. Cerebral white matter hyperintensities predict functional stroke outcome. Cerebrovasc Dis 29, 22–27 (2010). 10.1159/000255970

63 DeCarli, C., Fletcher, E., Ramey, V., Harvey, D. & Jagust, W. J. Anatomical mapping of white matter hyperintensities (WMH): exploring the relationships between periventricular WMH, deep WMH, and total WMH burden. Stroke 36, 50–55 (2005). 10.1161/01.STR.0000150668.58689.f2

64 Caligiuri, M. E. et al. Automatic Detection of White Matter Hyperintensities in Healthy Aging and Pathology Using Magnetic Resonance Imaging: A Review. Neuroinformatics 13, 261–276 (2015). 10.1007/s12021-015-9260-y

65 van Straaten, E. C. et al. Impact of white matter hyperintensities scoring method on correlations with clinical data: the LADIS study. Stroke 37, 836–840 (2006). 10.1161/01.STR.0000202585.26325.74

66 Medrano-Martorell, S. et al. Risk factors analysis according to regional distribution of white matter hyperintensities in a stroke cohort. Eur Radiol 32, 272–280 (2022). 10.1007/s00330-021-08106-2

67 Barkhof, F. & Scheltens, P. Is the whole brain periventricular? J Neurol Neurosurg Psychiatry 77, 143–144 (2006). 10.1136/jnnp.2005.075101

68 Giese, A. K. et al. White matter hyperintensity burden in acute stroke patients differs by ischemic stroke subtype. Neurology 95, e79–e88 (2020). 10.1212/WNL.0000000000009728

69 Rost, N. S. et al. White matter hyperintensity volume is increased in small vessel stroke subtypes. Neurology 75, 1670–1677 (2010). 10.1212/WNL.0b013e3181fc279a

70 de Leeuw, F. E. et al. Atrial fibrillation and the risk of cerebral white matter lesions. Neurology 54, 1795–1801 (2000). 10.1212/wnl.54.9.1795

71 Kim, S. et al. Periventricular white matter hyperintensities and the risk of dementia: a CREDOS study. Int Psychogeriatr 27, 2069–2077 (2015). 10.1017/S1041610215001076

72 Zheng, K. et al. Analysis of Risk Factors for White Matter Hyperintensity in Older Adults without Stroke. Brain Sci 13 (2023). 10.3390/brainsci13050835

73 Jimenez-Conde, J. et al. Hyperlipidemia and reduced white matter hyperintensity volume in patients with ischemic stroke. Stroke 41, 437–442 (2010). 10.1161/STROKEAHA.109.563502

74 Kalinin, M. N., Khasanova, D. R. & Ibatullin, M. M. The hemorrhagic transformation index score: a prediction tool in middle cerebral artery ischemic stroke. BMC Neurol 17, 177 (2017). 10.1186/s12883-017-0958-3

75 Melazzini, L. et al. White Matter Hyperintensities Quantification in Healthy Adults: A Systematic Review and Meta-Analysis. J Magn Reson Imaging 53, 1732–1743 (2021). 10.1002/jmri.27479

76 Grosu, S. et al. Associated factors of white matter hyperintensity volume: a machine-learning approach. Sci Rep 11, 2325 (2021). 10.1038/s41598-021-81883-4

77 Bjornfot, C. et al. Cerebral arterial stiffness is linked to white matter hyperintensities and perivascular spaces in older adults - A 4D flow MRI study. J Cereb Blood Flow Metab 44, 1343–1351 (2024). 10.1177/0271678X241230741

78 Wright, C. B. et al. Subclinical Cerebrovascular Disease Increases the Risk of Incident Stroke and Mortality: The Northern Manhattan Study. J Am Heart Assoc 6 (2017). 10.1161/JAHA.116.004069

79 Ghaznawi, R. et al. Association of White Matter Hyperintensity Markers on MRI and Long-term Risk of Mortality and Ischemic Stroke: The SMART-MR Study. Neurology 96, e2172–e2183 (2021). 10.1212/WNL.0000000000011827

80 Smirnov, M., Destrieux, C. & Maldonado, I. L. Cerebral white matter vasculature: still uncharted? Brain 144, 3561–3575 (2021). 10.1093/brain/awab273

81 Mok, V. et al. Race-ethnicity and cerebral small vessel disease--comparison between Chinese and White populations. Int J Stroke 9 Suppl A100, 36–42 (2014). 10.1111/ijs.12270

82 Ko, Y. et al. MRI-based Algorithm for Acute Ischemic Stroke Subtype Classification. J Stroke 16, 161–172 (2014). 10.5853/jos.2014.16.3.161

83 Adams, H. P., Jr., et al. Classification of subtype of acute ischemic stroke. Definitions for use in a multicenter clinical trial. TOAST. Trial of Org 10172 in Acute Stroke Treatment. Stroke 24, 35–41 (1993). 10.1161/01.str.24.1.35

84 van Swieten, J. C., Koudstaal, P. J., Visser, M. C., Schouten, H. J. & van Gijn, J. Interobserver agreement for the assessment of handicap in stroke patients. Stroke 19, 604–607 (1988). 10.1161/01.str.19.5.604

85 Brott, T. et al. Measurements of acute cerebral infarction: a clinical examination scale. Stroke 20, 864–870 (1989). 10.1161/01.str.20.7.864

86 Jeong, H. G., Kim, B. J., Yang, M. H., Han, M. K. & Bae, H. J. Neuroimaging markers for early neurologic deterioration in single small subcortical infarction. Stroke 46, 687–691 (2015). 10.1161/STROKEAHA.114.007466

87 Park, T. H. et al. Neurologic deterioration in patients with acute ischemic stroke or transient ischemic attack. Neurology 95, e2178–e2191 (2020). 10.1212/WNL.0000000000010603

88 Park, H. K. et al. One-Year Outcomes After Minor Stroke or High-Risk Transient Ischemic Attack: Korean Multicenter Stroke Registry Analysis. Stroke 48, 2991–2998 (2017). 10.1161/STROKEAHA.117.018045

89 Miller, K. L. et al. Multimodal population brain imaging in the UK Biobank prospective epidemiological study. Nat Neurosci 19, 1523–1536 (2016). 10.1038/nn.4393

90 Rorden, C. & Brett, M. Stereotaxic display of brain lesions. Behav Neurol 12, 191–200 (2000). 10.1155/2000/421719

91 Fischl, B. FreeSurfer. Neuroimage 62, 774–781 (2012). 10.1016/j.neuroimage.2012.01.021

92 Park, G., Hong, J., Duffy, B. A., Lee, J. M. & Kim, H. White matter hyperintensities segmentation using the ensemble U-Net with multi-scale highlighting foregrounds. Neuroimage 237, 118140 (2021). 10.1016/j.neuroimage.2021.118140

93 Kuijf, H. J. et al. Standardized Assessment of Automatic Segmentation of White Matter Hyperintensities and Results of the WMH Segmentation Challenge. IEEE Trans Med Imaging 38, 2556–2568 (2019). 10.1109/TMI.2019.2905770

94 Andersson, J. L. Non-linear registration, aka spatial normalisation, FMRIB technical report. FMRIB Anal Group Univ Oxford 2, e21 (2010).

95 Sanroma, G. et al. in Machine Learning in Medical Imaging: 9th International Workshop, MLMI 2018, Held in Conjunction with MICCAI 2018, Granada, Spain, September 16, 2018, Proceedings 9. 81–88 (Springer).

96 Fonteijn, H. M. et al. An event-based model for disease progression and its application in familial Alzheimer’s disease and Huntington’s disease. Neuroimage 60, 1880–1889 (2012). 10.1016/j.neuroimage.2012.01.062

97 Young, A. L. et al. A data-driven model of biomarker changes in sporadic Alzheimer’s disease. Brain 137, 2564–2577 (2014). 10.1093/brain/awu176

98 Mize, T. D., Doan, L. & Long, J. S. A general framework for comparing predictions and marginal effects across models. Sociological Methodology 49, 152–189 (2019).

99 DeLong, E. R., DeLong, D. M. & Clarke-Pearson, D. L. Comparing the areas under two or more correlated receiver operating characteristic curves: a nonparametric approach. Biometrics 44, 837–845 (1988).

100 Stuart, E. A., King, G., Imai, K. & Ho, D. MatchIt: nonparametric preprocessing for parametric causal inference. Journal of statistical software (2011).

101 Greifer, N. Covariate balance tables and plots: a guide to the cobalt package. Accessed March 10, 2020 (2020).

