## Supplementary Material for "Distinct Spatiotemporal Patterns of White Matter Hyperintensity Progression"

Jinyong Chung *et al.*

**Supplementary Information**

Supplementary Tables 1–12

Supplementary Figures 1–10

**Supplementary Tables**

**Supplementary Table 1 | Mixed-effects regression model for longitudinal subtype stability**

| **Longitudinal subtype stability (Unstable=0, Stable=1)** | | | | |
| --- | --- | --- | --- | --- |
| **Parameters** | | **OR** | **95% CI** | ***P*** |
| **WMH subtype**  **(vs. T2: radial)** | **T1: FP** | 0.887 | 0.385–2.043 | 0.777 |
|  | **T3: TO** | 0.871 | 0.291–2.608 | 0.804 |
| **FU years** | | 0.989 | 0.884–1.107 | 0.847 |
| **Age, year** | | 0.977 | 0.948–1.008 | 0.140 |
| **Sex (male=1)** | | 0.551 | 0.221–1.370 | 0.198 |
| **HTN** | | 1.021 | 0.472–2.206 | 0.958 |
| **DM** | | 0.426 | 0.222–0.816 | 0.010 |
| **HL** | | 0.962 | 0.494–1.873 | 0.908 |
| **Smoking** | | 1.602 | 0.664–3.864 | 0.293 |
| **AF** | | 1.213 | 0.441–3.336 | 0.708 |
| **CAD** | | 1.549 | 0.525–4.569 | 0.427 |
| **Revascularization therapy** | | 0.270 | 0.084–0.866 | 0.028 |

OR, odd ratio; CI, confidence interval; WMH, white matter hyperintensity; FP, fronto-parietal; TO, temporo-occipital; FU, follow-up; HTN, hypertension; DM, diabetes mellitus; HL, hyperlipidemia; AF, atrial fibrillation; CAD, coronary artery disease.

**Supplementary Table 2 | Mixed-effects regression models for demographics and vascular risk factors according to white matter hyperintensity subtype and stage**

| Parameter | | | **Age** | **Sex** | **HTN** | **DM** | **HL** | **Smoking** | **AF** | **CAD** |
| --- | --- | --- | --- | --- | --- | --- | --- | --- | --- | --- |
| **Subtype**  **(vs. T2: radial)** | **T1: FP** | Beta*/OR  (95% CI) | 9.35* (8.10‒10.59) | 0.89 (0.79‒1.00) | 1.47 (1.13‒1.91) | 0.93 (0.82‒1.05) | 1.01 (0.88‒1.15) | 0.98 (0.84‒1.14) | 0.92 (0.79‒1.06) | 1.05 (0.87‒1.28) |
|  |  | *T* | 14.68 | -1.94 | 2.89 | -1.19 | 0.08 | -0.25 | -1.17 | 0.52 |
|  |  | *P* | < 0.001 | 0.053 | 0.004 | 0.233 | 0.935 | 0.801 | 0.242 | 0.602 |
|  | **T3: TO** | Beta*/OR  (95% CI) | 0.00* (-1.23‒1.23) | 2.27 (2.03‒2.54) | 0.94 (0.74‒1.21) | 1.05 (0.04‒1.17) | 0.89 (0.78‒1.00) | 1.02 (0.90‒1.16) | 1.56 (1.37‒1.77) | 1.28 (1.07‒1.52) |
|  |  | *T* | 0.00 | 14.33 | -0.45 | 0.84 | -1.91 | 0.29 | 6.72 | 2.76 |
|  |  | *P* | 1.000 | < 0.001 | 0.651 | 0.399 | 0.056 | 0.770 | < 0.001 | 0.006 |
| **Stage** | | Beta*/OR  (95% CI) | 1.39* (1.32‒1.46) | 1.03 (1.02‒1.04) | 1.05 (1.03‒1.07) | 1.02 (1.01‒1.03) | 0.99 (0.98‒1.01) | 1.00 (0.98‒1.01) | 0.96 (0.95‒0.97) | 0.98 (0.96‒0.99) |
|  |  | *T* | 38.36 | 4.99 | 6.08 | 4.16 | -1.13 | -0.75 | -6.49 | -2.69 |
|  |  | *P* | < 0.001 | < 0.001 | < 0.001 | < 0.001 | 0.261 | 0.453 | < 0.001 | 0.007 |
| **Subtype–Stage interaction** | **T1*Stage** | Beta*/OR  (95% CI) | -0.77* (-0.88‒-0.66) | - | 0.98 (0.95‒1.00) | - | - | - | - | - |
|  |  | *T* | -13.76 | - | -2.03 | - | - | - | - | - |
|  |  | *P* | < 0.001 | - | 0.043 | - | - | - | - | - |
|  | **T3*Stage** | Beta*/OR  (95% CI) | -0.19* (-0.30‒-0.08) | - | 1.00 (0.98‒1.02) | - | - | - | - | - |
|  |  | *T* | -3.36 | - | -0.083 | - | - | - | - | - |
|  |  | *P* | < 0.001 | - | 0.933 | - | - | - | - | - |

Statistics (Beta coefficient, OR: odd ratio, *T*: t-statistic) for subtype and stage from mixed-effects regression models that included each variable along with age (except in the case of age), sex (except in the case of sex) as fixed effects, and imaging center as a random effect. HTN, hypertension; DM, diabetes mellitus; HL, hyperlipidemia; AF, atrial fibrillation; CAD, coronary artery disease; FP, fronto-parietal; TO, temporo-occipital; CI, confidence interval.

**Supplementary Table 3 | Mixed-effects regression model for age and hypertension in stroke patients with white matter hyperintensity stage ≤ 9**

|  |  | **Age, year** |  | | **Hypertension, %** |  | |
| --- | --- | --- | --- | --- | --- | --- | --- |
| **Parameter** |  | **Beta (95% CI)** | | ***P*** | **OR (95% CI)** | | ***P*** |
| **Subtype**  **(vs. T2: radial)** | **T1: FP** | 5.87 (4.90‒6.84) | | < 0.001 | 1.31 (1.08‒1.59) | | 0.005 |
|  | **T3: TO** | -2.56 (-3.41‒-1.72) | | < 0.001 | 0.91 (0.78‒1.07) | | 0.254 |
| **Stage** |  | 1.95 (1.82‒2.08) | | < 0.001 | 1.07 (1.04‒1.10) | | < 0.001 |
| **Age** |  | - | | - | 1.03 (1.02‒1.03) | | < 0.001 |
| **Sex** |  | -5.85 (-6.54‒-5.16) | | < 0.001 | 0.83 (0.73‒0.95) | | 0.008 |

CI, confidence interval; OR, odd ratio; FP, fronto-parietal; TO, temporo-occipital.

**Supplementary Table 4 | Mixed-effects regression model for age and hypertension in stroke patients with white matter hyperintensity stage > 9**

|  |  | **Age, year** |  | | **Hypertension, %** |  | |
| --- | --- | --- | --- | --- | --- | --- | --- |
| **Parameter** |  | **Beta (95% CI)** | | ***P*** | **OR (95% CI)** | | ***P*** |
| **Subtype**  **(vs. T2: radial)** | **T1: FP** | -0.55 (-1.31‒0.21) | | 0.157 | 1.07 (0.88‒1.29) | | 0.505 |
|  | **T3: TO** | -1.55 (-2.24‒-0.87) | | < 0.001 | 0.93 (0.79‒1.10) | | 0.378 |
| **Stage** |  | 0.58 (0.48‒0.68) | | < 0.001 | 1.02 (0.99‒1.05) | | 0.125 |
| **Age** |  | - | | - | 1.01 (1.00‒1.01) | | 0.135 |
| **Sex** |  | -5.65 (-6.23‒-5.07) | | < 0.001 | 0.69 (0.59‒0.80) | | < 0.001 |

CI, confidence interval; OR, odd ratio; FP, fronto-parietal; TO, temporo-occipital.

**Supplementary Table 5 | Demographics, vascular risk factors, and stroke etiologies by subtype and stage of white matter hyperintensity progression in stroke patients without adjustments for age and sex**

|  | **T1: FP** | **T2: radial** | **T3: TO** | ***P*^*^** | | |
| --- | --- | --- | --- | --- | --- | --- |
| ***n* (% of total)** | 1870 (21.4) | 4014 (46.0) | 2852 (32.7) | **subtype** | **stage** | **interaction** |
| **Age, year (SD)** | 72.4 (10.8) | 66.2 (13.1) | 67.9 (12.0) | < 0.001 | < 0.001 | < 0.001 |
| **Sex, male %** | 46.7 | 54.7 | 72.3 | < 0.001 | < 0.001 | - |
| **HTN, %** | 73.2 | 62.7 | 65.3 | < 0.001 | < 0.001 | - |
| **DM, %** | 30.1 | 29.8 | 32.8 | 0.081 | < 0.001 | - |
| **HL, %** | 28.6 | 27.8 | 26.5 | 0.032 | 0.012 | - |
| **Smoking, %** | 31.8 | 38.9 | 47.6 | < 0.001 | < 0.001 | - |
| **AF, %** | 19.8 | 18.5 | 24.2 | < 0.001 | < 0.001 | - |
| **CAD, %** | 10.7 | 8.9 | 11.4 | 0.042 | 0.068 | - |
| **SBI, %** | 58.4 | 34.2 | 40.1 | < 0.001 | < 0.001 | - |
| **CMB, %** | 30.0 | 14.8 | 20.4 | < 0.001 | < 0.001 | - |
| **Stroke subtypes** |  |  |  |  |  |  |
| LAA, % | 38.6 | 38.5 | 36.7 | 0.311 | 0.665 | - |
| SVO, % | 16.7 | 17.5 | 15.0 | < 0.001 | 0.122 | - |
| CE, % | 20.7 | 20.3 | 24.5 | 0.001 | 0.568 | - |
| **Acute infarct volume (IQR), ml** | 2.46  (0.64‒13.77) | 2.36  (0.57‒13.14) | 3.53  (0.60‒18.21) | < 0.001 | < 0.001 | - |
| **Admission**  **NIHSS score (IQR)** | 4 (2‒9) | 3 (1‒8) | 4 (2‒9) | < 0.001 | < 0.001 | - |

**P* from mixed-effects regression models that included each variable along with subtype, stage, and the subtype–stage interaction as fixed effects, and imaging center as a random effect. Onset to MRI time was added as a fixed factor for acute infarct volume. FP, fronto-parietal; TO, temporo-occipital; SD, standard deviation; HTN, hypertension; DM, diabetes mellitus; HL, hyperlipidemia; AF, atrial fibrillation; CAD, coronary artery disease; SBI, silent brain infarct; CMB, cerebral microbleeds; LAA, large artery atherosclerosis; SVO, small vessel occlusion; CE, cardioembolism; IQR, interquartile range; NIHSS, National Institute of Health Stroke Scale.

**Supplementary Table 6 | Mixed-effects regression models for stroke etiologies according to white matter hyperintensity subtype and stage**

**in stroke patients**

| Parameter | | | **SBI presence** | **CMB presence** | **Stroke subtype (vs. others)** | | | **Infarct volume** | **Admission NIHSS score** |
| --- | --- | --- | --- | --- | --- | --- | --- | --- | --- |
|  |  |  |  |  | **LAA** | **SVO** | **CE** |  |  |
| **Subtype**  **(vs. T2: radial)** | **T1: FP** | Beta*/OR  (95% CI) | 1.38 (1.20‒1.58) | 1.37 (1.18‒1.59) | 1.04 (0.92‒1.17) | 0.89 (0.76‒1.04) | 0.93 (0.80‒1.07) | 0.18* (0.06‒0.29) | 0.05* (-0.01‒0.10) |
|  |  | *T* | 4.65 | 4.19 | 0.57 | -1.46 | -1.04 | 2.90 | 1.66 |
|  |  | *P* | < 0.001 | < 0.001 | 0.570 | 0.145 | 0.300 | 0.004 | 0.098 |
|  | **T3: TO** | Beta*/OR  (95% CI) | 0.56 (0.49‒0.63) | 0.83 (0.72‒0.95) | 0.91 (0.81‒1.01) | 0.68 (0.59‒0.79) | 1.45 (1.28‒1.65) | 0.35* (0.25‒0.46) | 0.17* (0.12‒0.22) |
|  |  | *T* | -9.58 | -2.61 | -1.81 | -5.23 | 5.85 | 6.34 | 6.53 |
|  |  | *P* | < 0.001 | 0.009 | 0.071 | < 0.001 | < 0.001 | < 0.001 | < 0.001 |
| **Stage** | | Beta*/OR  (95% CI) | 1.24 (1.22‒1.25) | 1.17 (1.15‒1.19) | 1.01 (1.00‒1.02) | 1.06 (1.04‒1.07) | 0.96 (0.95‒0.97) | -0.05* (-0.06‒-0.04) | -0.01* (-0.01‒-0.00) |
|  |  | *T* | 33.28 | 22.69 | 1.11 | 8.08 | -7.09 | -9.87 | -3.68 |
|  |  | *P* | < 0.001 | < 0.001 | 0.269 | < 0.001 | < 0.001 | < 0.001 | < 0.001 |

Statistics (Beta coefficient, OR: odd ratio, *T*: t-statistic) for subtype and stage from mixed-effects regression models that included each variable along with age (except in the case of age), sex (except in the case of sex) as fixed effects, and imaging center as a random effect. Onset to MRI time was added as a fixed factor for acute infarct volume. SBI, silent brain infarct; CMB, cerebral microbleeds; LAA, large artery atherosclerosis; SVO, small vessel occlusion; CE, cardioembolism; NIHSS, National Institute of Health Stroke Scale; FP, fronto-parietal; TO, temporo-occipital; CI, confidence interval.

**Supplementary Table 7 | Mixed-effects regression models for demographics and vascular risk factors according to Fazekas scale in stroke patients**

| Parameter | | **Age** | **Sex** | **HTN** | **DM** | **HL** | **Smoking** | **AF** | **CAD** |
| --- | --- | --- | --- | --- | --- | --- | --- | --- | --- |
| **Total Fazekas scale (0-6)** | Beta*/OR  (95% CI) | 3.65* (3.50‒3.80) | 0.93 (0.90‒0.96) | 1.16 (1.12‒1.20) | 1.05 (1.02‒1.09) | 1.03 (1.00‒1.07) | 1.02 (0.98‒1.06) | 0.87 (0.83‒0.90) | 1.02 (0.96‒1.07) |
|  | *T* | 48.29 | -4.53 | 8.46 | 3.06 | 1.90 | 0.91 | -6.89 | 0.59 |
|  | *P* | < 0.001 | < 0.001 | < 0.001 | 0.002 | 0.058 | 0.361 | < 0.001 | 0.555 |
| **PVWMH Fazekas scale (0-3)** | Beta*/OR  (95% CI) | 4.49* (4.09‒4.88) | 0.92 (0.85‒0.99) | 1.13 (1.04‒1.22) | 1.10 (1.01‒1.19) | 1.08 (0.99‒1.17) | 0.95 (0.86‒1.04) | 0.74 (0.67‒0.82) | 1.00 (0.88‒1.14) |
|  | *T* | 22.18 | -2.15 | 2.88 | 2.24 | 1.74 | -1.11 | -5.98 | 0.01 |
|  | *P* | < 0.001 | 0.032 | 0.004 | 0.025 | 0.083 | 0.269 | < 0.001 | 0.994 |
| **DWMH Fazekas scale (0-3)** | Beta*/OR  (95% CI) | 2.91* (2.55‒3.26) | 0.94 (0.87‒1.01) | 1.18 (1.10‒1.27) | 1.02 (0.94‒1.09) | 1.00 (0.92‒1.08) | 1.08 (1.00‒1.18) | 1.00 (0.91‒1.09) | 1.03 (1.79‒2.12) |
|  | *T* | 15.86 | -1.75 | 4.54 | 0.43 | -0.07 | 1.86 | -0.09 | 0.49 |
|  | *P* | < 0.001 | 0.081 | < 0.001 | 0.669 | 0.945 | 0.062 | 0.926 | 0.622 |

Statistics (Beta coefficient, OR: odd ratio, *T*: t-statistic) for Fazekas scales from mixed-effects regression models that included each variable along with age (except in the case of age) and sex (except in the case of sex) as fixed effects, and imaging center as a random effect. HTN, hypertension; DM, diabetes mellitus; HL, hyperlipidemia; AF, atrial fibrillation; CAD, coronary artery disease; PVWMH, periventricular white matter hyperintensity; DWMH, deep white matter hyperintensity; CI, confidence interval.

**Supplementary Table 8 | Mixed-effects regression models for stroke etiologies according to Fazekas scale in stroke patients**

| Parameter | | **SBI presence** | **CMB presence** | **Stroke subtype (vs. others)** | | | **Infarct volume** | **Admission NIHSS score** |
| --- | --- | --- | --- | --- | --- | --- | --- | --- |
|  |  |  |  | **LAA** | **SVO** | **CE** |  |  |
| **Total Fazekas scale (0-6)** | Beta*/OR  (95% CI) | 1.86 (1.80‒1.94) | 1.67 (1.60‒1.74) | 1.02 (0.98‒1.05) | 1.21 (1.16‒1.26) | 0.87 (0.84‒0.91) | -0.12* (-0.30‒-0.11) | -0.02* (-0.04‒-0.01) |
|  | *T* | 32.48 | 23.99 | 0.98 | 8.80 | -6.91 | -6.63 | -3.24 |
|  | *P* | < 0.001 | < 0.001 | 0.328 | < 0.001 | < 0.001 | < 0.001 | 0.001 |
| **PVWMH Fazekas scale (0-3)** | Beta*/OR  (95% CI) | 1.95 (1.79‒2.12) | 1.84 (1.66‒2.04) | 1.04 (0.96‒1.13) | 1.33 (1.20‒1.47) | 0.78 (0.71‒0.86) | -0.26* (-0.34‒-0.17) | -0.05* (-0.08‒-0.00) |
|  | *T* | 15.39 | 11.78 | 1.04 | 5.34 | -5.22 | -6.18 | -2.42 |
|  | *P* | < 0.001 | < 0.001 | 0.297 | < 0.001 | < 0.001 | < 0.001 | 0.015 |
| **DWMH Fazekas scale (0-3)** | Beta*/OR  (95% CI) | 1.79 (1.66‒1.93) | 1.54 (1.40‒1.68) | 0.99 (0.93‒1.07) | 1.11 (1.02‒1.22) | 0.96 (0.89‒1.05) | 0.01* (-0.06‒0.08) | -0.01* (-0.04‒0.03) |
|  | *T* | 15.09 | 9.16 | -0.18 | 2.29 | -0.88 | 0.21 | -0.40 |
|  | *P* | < 0.001 | < 0.001 | 0.857 | 0.022 | 0.379 | 0.835 | 0.692 |

Statistics (Beta coefficient, OR: odd ratio, *T*: t-statistic) for Fazekas scales from mixed-effects regression models that included each variable along with age (except in the case of age), sex (except in the case of sex) as fixed effects, and imaging center as a random effect. Onset to MRI time was added as a fixed factor for acute infarct volume. SBI, silent brain infarct; CMB, cerebral microbleeds; LAA, large artery atherosclerosis; SVO, small vessel occlusion; CE, cardioembolism; NIHSS, National Institute of Health Stroke Scale; PVWMH, periventricular white matter hyperintensity; DWMH, deep white matter hyperintensity; CI, confidence interval.

**Supplementary Table 9 | Mixed-effects and cox proportional hazards regression models for post-stroke outcomes according to Fazekas scale in stroke patients**

| Parameter | | **Early (< 3 weeks) neurological deterioration** | | | | **3-month** | **1-year stroke recurrence** | | **1-year mortality** | | |
| --- | --- | --- | --- | --- | --- | --- | --- | --- | --- | --- | --- |
|  |  | Total | Stroke recurrence | Stroke progression | Symptomatic  HT | mRS score > 3 | Ischemic | Hemorrhagic | All-cause | Vascular | Nonvascular |
| **Total Fazekas scale (0-6)** | OR/HR* (95% CI) | 1.06 (1.01‒1.10) | 1.08 (0.94‒1.25) | 1.05 (1.00‒1.10) | 0.80 (0.68‒0.94) | 1.04 (1.00‒1.09) | 1.16* (0.99‒1.37) | 1.82* (1.11‒2.97) | 1.08* (1.07‒1.09) | 1.14* (0.80‒1.62) | 1.14* (1.02‒1.28) |
|  | *T/Z** | 2.41 | 1.12 | 1.98 | -2.75 | 1.89 | 1.85* | 2.37* | 2.49* | 0.71* | 2.39* |
|  | *P* | 0.011 | 0.263 | 0.047 | 0.006 | 0.059 | 0.064 | 0.018 | 0.013 | 0.480 | 0.017 |
| **PVWMH Fazekas scale (0-3)** | OR/HR* (95% CI) | 1.05 (0.95‒1.16) | 0.90 (0.64‒1.28) | 1.09 (0.97‒1.22) | 0.57 (0.39‒0.82) | 1.08 (0.97‒1.20) | 1.04 (0.84‒1.27) | 1.61* (0.84‒3.08) | 1.06* (0.93‒1.22) | 1.17* (0.74‒1.85) | 1.05* (0.91‒1.22) |
|  | *T/Z** | 0.80 | -0.59 | 1.45 | -3.00 | 1.46 | 0.35* | 1.44* | 0.89* | 0.65* | 0.73* |
|  | *P* | 0.351 | 0.556 | 0.15 | 0.003 | 0.145 | 0.729 | 0.149 | 0.376 | 0.513 | 0.465 |
| **DWMH Fazekas scale (0-3)** | OR/HR* (95% CI) | 1.06 (0.97‒1.16) | 1.28 (0.93‒1.75) | 1.02 (0.92‒1.12) | 1.07 (0.77‒1.48) | 1.01 (0.92‒1.11) | 1.18 (0.98‒1.42) | 1.18* (0.66‒2.11) | 1.11* (0.98‒1.25) | 0.97* (0.64‒1.45) | 1.12* (0.99‒1.28) |
|  | *T/Z** | 1.28 | 1.52 | 0.29 | 0.40 | 0.20 | 1.73* | 0.54* | 1.64* | -0.17* | 1.77* |
|  | *P* | 0.208 | 0.128 | 0.774 | 0.694 | 0.843 | 0.084 | 0.586 | 0.101 | 0.865 | 0.077 |

Statistics (OR: odd ratio, HR: hazard ratio, *T*: t-statistic, *Z*: z-statistic) for Fazekas scales from mixed-effects regression models that included each variable along with age (except in the case of age), sex (except in the case of sex), and revascularization therapy as fixed effects, plus imaging center as a random effect. For 3-month unfavorable functional outcome (modified Rankin Scale [mRS] score > 3), only patients with pre-stroke mRS scores < 2 were included, and pre-stroke mRS score was added as a covariate. HT, hemorrhagic transformation; PVWMH, periventricular white matter hyperintensity; DWMH, deep white matter hyperintensity; CI, confidence interval.

**Supplementary Table 10 | Demographics and vascular risk factors by subtype and stage of white matter hyperintensity progression in high-risk controls**

|  | **T1: FP** | **T2: radial** | **T3: TO** | ***P*^*^** | | |
| --- | --- | --- | --- | --- | --- | --- |
| ***n* (% of total)** | 4558 (45.7) | 4155 (41.7) | 1254 (12.6) | **subtype** | **stage** | **interaction** |
| **Age, year (SD)** | 68.4 (6.0) | 67.7 (6.6) | 65.3 (6.9) | < 0.001 | < 0.001 | < 0.001 |
| **Sex, male %** | 47.9 | 50.9 | 56.9 | < 0.001 | 0.550 | - |
| **HTN, %** | 59.2 | 55.5 | 52.2 | 0.007 | < 0.001 | - |
| **DM, %** | 10.6 | 10.3 | 9.4 | 0.226 | 0.015 | - |
| **HL, %** | 32.5 | 31.7 | 30.6 | 0.945 | 0.021 | - |
| **Smoking, %** | 58.0 | 58.0 | 59.0 | 0.902 | <0.001 | - |
| **AF, %** | 7.4 | 7.2 | 7.2 | 0.771 | 0.437 | - |
| **CAD, %** | 13.2 | 12.4 | 11.7 | 0.513 | 0.221 | - |

**P* from mixed-effects regression models that included each variable along with age (except in the case of age), sex (except in the case of sex), subtype, stage, and subtype–stage interaction as fixed effects, with imaging center as a random effect. Onset to MRI time was added as a fixed factor for acute infarct volume. FP, fronto-parietal; TO, temporo-occipital; SD, standard deviation; HTN, hypertension; DM, diabetes mellitus; HL, hyperlipidemia; AF, atrial fibrillation; CAD, coronary artery disease.

**Supplementary Table 11 | Demographics, vascular risk factors, and stroke etiologies by subtype and stage of white matter hyperintensity progression in stroke patients age- and sex-matched to low-risk controls**

|  | **T1: FP** | **T2: radial** | **T3: TO** | ***P*^*^** | | |
| --- | --- | --- | --- | --- | --- | --- |
| ***n* (% of total)** | 1670 (24.2) | 3180 (46.1) | 2049 (29.7) | **subtype** | **stage** | **interaction** |
| **Age, year (SD)** | 69.4 (9.1) | 63.9 (10.9) | 64.4 (10.2) | < 0.001 | < 0.001 | < 0.001 |
| **Sex, male %** | 46.5 | 58.1 | 69.9 | < 0.001 | < 0.001 | - |
| **HTN, %** | 73.1 | 62.0 | 64.2 | 0.013 | 0.003 | 0.105 |
| **DM, %** | 31.7 | 31.4 | 34.9 | 0.020 | < 0.001 | - |
| **HL, %** | 29.1 | 29.1 | 28.1 | 0.726 | 0.210 | - |
| **Smoking, %** | 33.7 | 42.5 | 49.3 | 0.836 | 0.231 | - |
| **AF, %** | 18.7 | 16.8 | 21.9 | < 0.001 | < 0.001 | - |
| **CAD, %** | 10.0 | 8.4 | 10.3 | 0.071 | 0.028 | - |
| **SBI, %** | 57.8 | 31.1 | 34.7 | < 0.001 | < 0.001 | - |
| **CMB, %** | 30.7 | 13.7 | 16.1 | < 0.001 | < 0.001 | - |
| **Stroke subtypes** |  |  |  |  |  |  |
| LAA, % | 40.1 | 38.7 | 37.4 | 0.108 | 0.829 | - |
| SVO, % | 19.2 | 19.2 | 16.2 | < 0.001 | < 0.001 | - |
| CE, % | 19.6 | 18.6 | 22.7 | 0.005 | 0.142 | < 0.001 |
| **Acute infarct volume (IQR), ml** | 2.25  (0.61‒12.70) | 2.15  (0.52‒11.96) | 3.22  (0.56‒17.78) | < 0.001 | < 0.001 | - |
| **Admission**  **NIHSS score (IQR)** | 4 (2‒8) | 3 (1‒7) | 4 (2‒8) | < 0.001 | 0.028 | - |

**P* from mixed-effects regression models that included each variable along with age (except in the case of age), sex (except in the case of sex), subtype, stage, and the subtype-stage interaction as fixed effects, with imaging center as a random effect. Onset to MRI time was added as a fixed factor for acute infarct volume. FP, fronto-parietal; TO, temporo-occipital; SD, standard deviation; HTN, hypertension; DM, diabetes mellitus; HL, hyperlipidemia; AF, atrial fibrillation; CAD, coronary artery disease; SBI, silent brain infarct; CMB, cerebral microbleeds; LAA, large artery atherosclerosis; SVO, small vessel occlusion; CE, cardioembolism; IQR, interquartile range; NIHSS, National Institute of Health Stroke Scale.

**Supplementary Table 12 | Mixed-effects and cox proportional hazards regression models for post-stroke outcomes according to white matter hyperintensity subtype and stage in stroke patients age- and sex-matched to low-risk controls**

| Parameter | | | **Early (< 3 weeks) neurological deterioration** | | | | **3-month** | **1-year stroke recurrence** | | **1-year mortality** | | |
| --- | --- | --- | --- | --- | --- | --- | --- | --- | --- | --- | --- | --- |
|  |  |  | Total | Stroke recurrence | Stroke progression | Symptomatic HT | mRS score > 3 | Ischemic | Hemorrhagic | All-cause | Vascular | Nonvascular |
| **Subtype**  **(vs. T2: radial)** | **T1: FP** | OR/HR* (95% CI) | 1.07 (0.90‒1.27) | 1.66 (0.93‒2.99) | 1.05 (0.87‒1.26) | 0.88 (0.44‒1.77) | 1.15 (0.96‒1.38) | 1.55* (1.10‒2.17) | 0.74* (0.25‒2.14) | 0.78* (0.60‒1.02) | 1.34* (0.60‒2.96) | 0.74* (0.56‒0.98) |
|  |  | *T/Z** | 0.79 | 1.71 | 0.46 | -0.36 | 1.53 | 2.53* | -0.56* | -1.79* | 0.72* | -2.12* |
|  |  | *P* | 0.430 | 0.087 | 0.643 | 0.721 | 0.126 | 0.011 | 0.572 | 0.073 | 0.473 | 0.034 |
|  | **T3: TO** | OR/HR* (95% CI) | 1.07 (0.92‒1.26) | 1.61 (0.91‒2.84) | 1.03 (0.87‒1.26) | 1.99 (1.16‒3.43) | 1.35 (1.14‒1.60) | 1.27* (0.91‒1.77) | 0.84* (0.32‒2.23) | 1.03* (0.81‒1.32) | 0.85* (0.36‒1.98) | 1.05* (0.82‒1.36) |
|  |  | *T/Z** | 0.87 | 1.65 | 0.35 | 2.49 | 3.51 | 1.43* | -0.35* | 0.27* | -0.39* | 0.39* |
|  |  | *P* | 0.384 | 0.099 | 0.729 | 0.012 | < 0.001 | 0.153 | 0.729 | 0.788 | 0.699 | 0.697 |
| **Stage** | | OR/HR* (95% CI) | 1.02 (1.00‒1.03) | 0.98 (0.93‒1.03) | 1.02 (1.00‒1.04) | 0.92 (0.87‒0.97) | 1.02 (1.00‒1.03) | 1.00* (0.97‒1.03) | 1.19* (1.08‒1.31) | 1.05* (1.02‒1.07) | 1.00* (0.93‒1.07) | 1.05* (1.03‒1.08) |
|  |  | *T/Z** | 2.28 | -0.85 | 2.37 | -2.91 | 2.21 | -0.09* | 3.60* | 3.89* | 0.06* | 4.09* |
|  |  | *P* | 0.023 | 0.394 | 0.018 | 0.004 | 0.027 | 0.927 | < 0.001 | < 0.001 | 0.950 | < 0.001 |

Statistics (OR: odd ratio, HR: hazard ratio, *T*: t-statistic, *Z*: z-statistic) for subtype and stage from mixed-effects regression models that included each variable along with age (except in the case of age), sex (except in the case of sex), and revascularization therapy as fixed effects, with imaging center as a random effect. For 3-month unfavorable functional outcome (modified Rankin Scale [mRS] score > 3), only patients with pre-stroke mRS scores < 2 were included, with the pre-stroke mRS score added as a covariate. HT, hemorrhagic transformation; CI, confidence interval..

**Supplementary Figure Legends**

**Supplementary Fig. 1 | Flow diagram.**

Diagram showing the inclusion and exclusion criteria of the UK Biobank and Korean MRI-based stroke database used in the study. FLAIR, fluid-attenuated inversion recovery; FU, follow-up.

**Supplementary Fig. 2 | Functional lobes-based bullseye parcellation of cerebral white matter.**

**a**, Five lobes (frontal: magenta, basal ganglia: yellow, temporal: blue, parietal: red, occipital: green) were each parcellated into four layers based on their relative distance to lateral ventricles and cortical surfaces (cyan gradients). **b**, 20 regions of interest were delineated to assess local white matter hyperintensity severity. The z-axis coordinates in the Montreal Neurological Institute space are displayed in the bottom right corner of the first three brain slices in the T1 group. R, right; L, left.

**Supplementary Fig. 3 | Cross-validation (CV) for evaluating different WMH subtyping–staging models.**

CV evaluation metrices across the number of subtypes in the functional lobe model **(a)** and the arterial territory model **(b)**.

**Supplementary Fig. 4 | Voxel-based white matter hyperintensity (WMH) frequency maps across distinct spatiotemporal trajectories of WMH progression in stroke patients.**

Voxel-based WMH frequency maps of distinct WMH progression subtypes (T1: fronto-parietal [FP]; T2: radial; T3: temporo-occipital [TO]). Bullseye presentations (F: frontal; B: basal ganglia; T: temporal; P: parietal; O: occipital) and Voxel-based WMH frequency maps for each WMH subtype at WMH stages 2, 6, 10, and 14. The z-axis coordinates in the Montreal Neurological Institute space are displayed in the bottom right corner of the first three brain slices in the T1 group.

**Supplementary Fig. 5 | Distinct demographics, risk factor, and etiology profiles of stroke patients with different white matter hyperintensity subtypes** **without adjustments for age and sex.**

Graphs of estimated marginal means (EMMs; a curve line with a shaded 95% confidence interval) for demographics, vascular risk factors, and etiologies across WMH stages within each WMH subtype (T1: fronto-parietal [FP]; T2: radial; T3: temporo-occipital [TO]) in stroke patients. Scatter plots of raw means were superimposed. The graph explicit presents significant inter-subtype differences across all stages. For factors showing significant subtype–stage interactions, we further stratified the stages into early and late periods based on median stage (9 for stroke patients), thereby defining inter-subtype differences across these periods. **a**, Age, hypertension (HTN), diabetes mellitus (DM), hyperlipidemia (HL), and current smoking. **b**, Atrial fibrillation (AF) and coronary artery disease (CAD). **c**, Presence of silent brain infarcts (SBIs) and cerebral microbleeds (CMBs). **d**, Stroke subtypes: large artery atherosclerosis (LAA), small vessel occlusion (SVO), and cardioembolism (CE). **e**, Acute infarct volume and admission National Institute of Health Stroke Scale (NIHSS) score.

**Supplementary Fig. 6 | Proportions of atrial fibrillation (AF) and coronary artery disease (CAD) by subtype and stage of white matter hyperintensity (WMH) progression in high-risk controls.**

Graphs of estimated marginal means (EMMs; a curved line with shaded 95% confidence interval) for proportions of AF and CAD in each WMH subtype (T1: fronto-parietal [FP]; T2: radial; T3: temporo-occipital [TO]). Scatter plots of raw means were superimposed.

**Supplementary Fig. 7 | Comparing white matter hyperintensity (WMH) stage and volume in order to discriminate stroke patients from high-risk controls.**

**a**, Distributions of WMH stages and volumes in high-risk controls and stroke patients. *P* values were calculated using Mann-Whitney *U*-tests to compare WMH stages and volumes between the groups. **b**, Receiver operating characteristic curves for distinguishing stroke patients from high-risk controls based on either WMH stage or WMH volume. Areas under the curves (AUCs) were calculated and statistically compared using the DeLong test.

**Supplementary Fig. 8 | Arterial territories-based bullseye parcellation of cerebral white matter.**

**a**, Each arterial territory (anterior cerebral artery: magenta, anterior cerebral artery-middle cerebral artery border zones: yellow, middle cerebral artery: blue, middle cerebral artery-posterior cerebral artery border zones: red, posterior cerebral artery: green) was parcellated into four layers based on its relative distance to lateral ventricles and cortical surfaces (cyan gradients). **b**, 20 regions of interest were finally delineated to assess local white matter hyperintensity severity. The z-axis coordinates in the Montreal Neurological Institute space are displayed in the bottom right corner of the first three brain slices in the T1 group. R, right; L, left.

**Supplementary Fig. 9 | Distinct spatiotemporal trajectories of white matter hyperintensity (WMH) progression in stroke patients matched to low-risk controls by age and sex.**

**a**, Spatiotemporal patterns of distinct WMH progression subtypes (T1: fronto-parietal [FP]; T2: radial; T3: temporo-occipital [TO]). Bullseye presentations (F: frontal; B: basal ganglia; T: temporal; P: parietal; O: occipital) and median WMH severity maps are presented for each WMH subtype at WMH stages 2, 6, 10, and 14. The z-axis coordinates in the Montreal Neurological Institute space are displayed in the bottom right corner of the first three brain slices in the T1 group. **b**, Left, Sankey diagram illustrating distributions of patients across the number of subtypes in Subtype and Stage Inference (SuStaIn) modeling. Groups in models were assigned the same color as the subtype with the highest proportion of patients based on the three-subtype model. Middle, distributions of stages across the subtypes. Patients at stage 0 were not included in any other subtypes (T1~T3). Right, scatter plot showing maximum likelihood (ML) subtype probabilities (prob.) of individuals in a 2D projection on a triangular plane. Graphs of estimated marginal means (EMMs; a curved line with shaded 95% confidence interval) for demographics, vascular risk factors, and stroke outcomes by stage in each subtype are shown **(c-g)**. Scatter plots of raw means were superimposed. The graph explicitly presents significant inter-subtype differences across all stages. Inter-subtype differences within stage 1–9 and 10–20 were also presented for factors showing significant subtype–stage interactions. **c**, Age. **d**, Hypertension (HTN) and atrial fibrillation (AF). **e**, Presence of silent brain infarcts (SBIs) and cerebral microbleeds (CMBs). **f**, Stroke subtypes: large artery atherosclerosis (LAA), small vessel occlusion (SVO), and cardioembolism (CE). **g**, Acute infarct volume and admission National Institute of Health Stroke Scale (NIHSS) score.

**Supplementary Fig. 10 | Post-stroke outcomes by subtype and stage of white matter hyperintensity (WMH) progression in stroke patients matched to low-risk controls by age and sex.**

Graphs of estimated marginal means (EMMs; a curved line with shaded 95% confidence interval) for post-stroke outcomes by WMH stage in each WMH subtype (T1: fronto-parietal [FP]; T2: radial; T3: temporo-occipital [TO]). Scatter plots of raw means were superimposed. **a**, Early (< 3 weeks) neurological deterioration and its causes (stroke recurrence, stroke progression, and symptomatic hemorrhagic transformation [HT]). **b**, Unfavorable functional outcome at 3 months (modified Rankin Scale score > 3). **c**, Stroke recurrence (ischemic and hemorrhagic) and mortality (all-cause and nonvascular death) within 1 year. Hazard ratio (HR) and *P* for stage are explicitly presented. Note that mortalities for T2 (black) and T3 (blue) are very similar, with closely overlapping curves.
