## Supplementary Figure for "Distinct Spatiotemporal Patterns of White Matter Hyperintensity Progression"

### Supplementary Figure 1 | Flow diagram of the study.

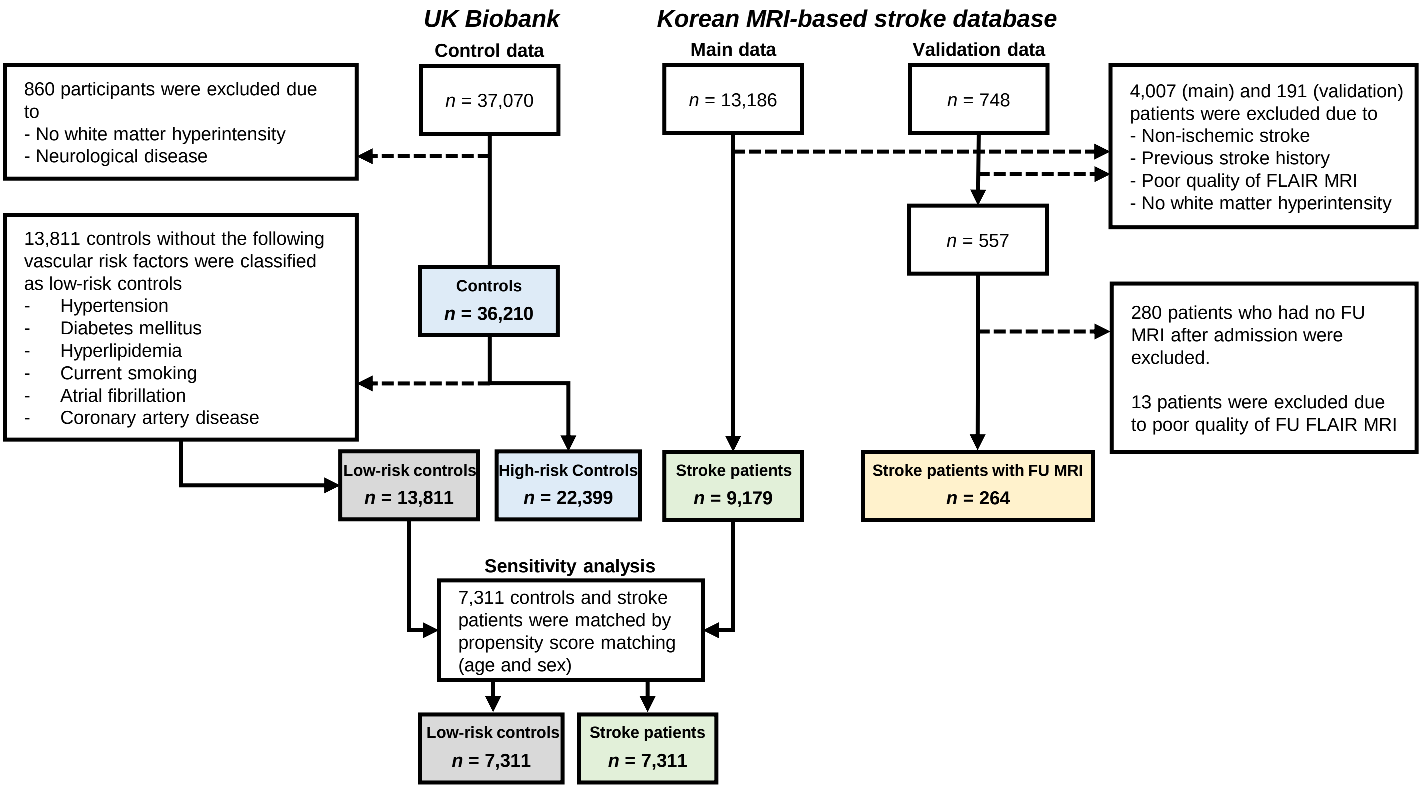

**Supplementary Figure 2 | Bullseye parcellation of cerebral white matter based on functional lobes.**

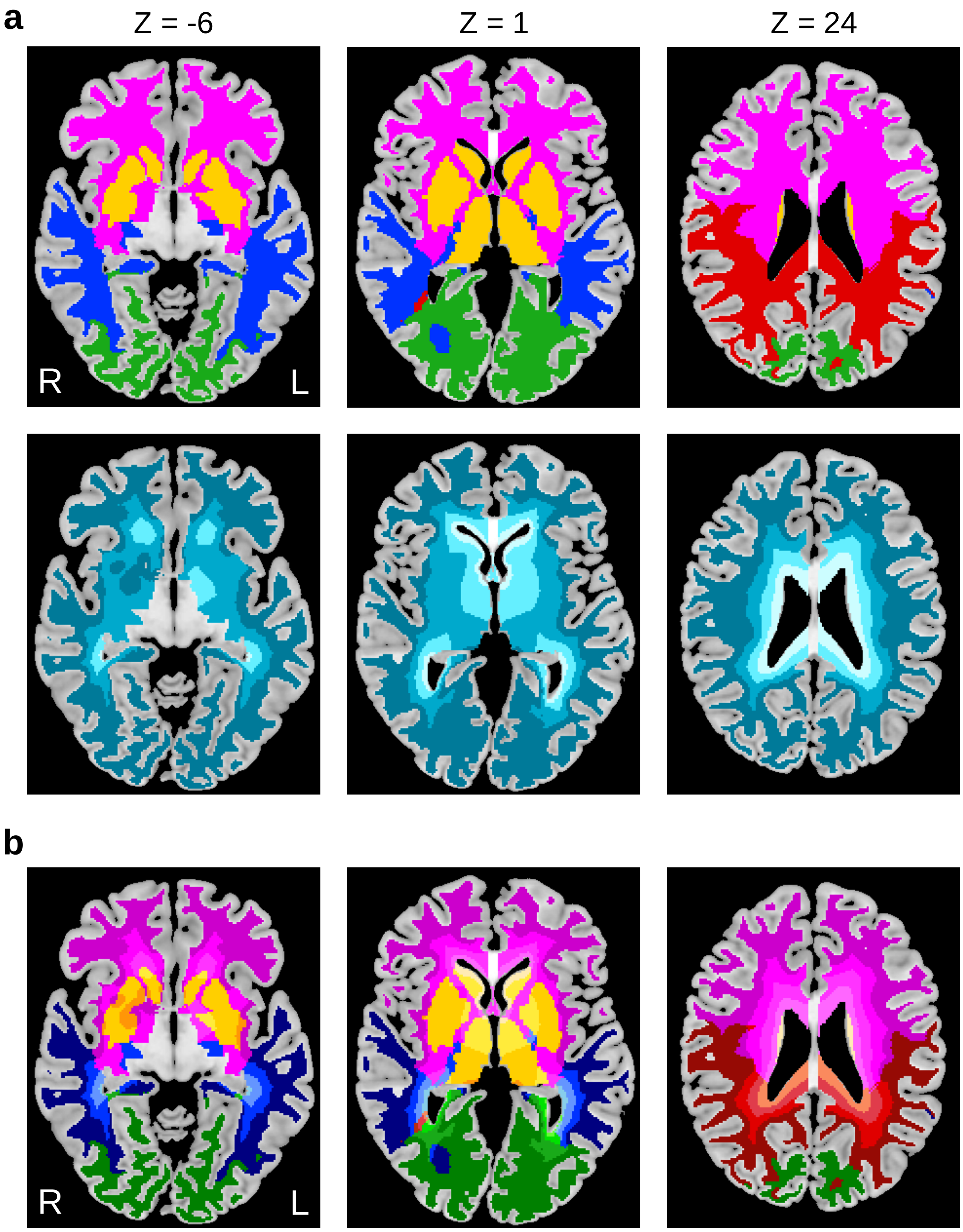

**Supplementary Figure 3 | Cross-validation for the evaluation of different WMH subtyping-staging models.**

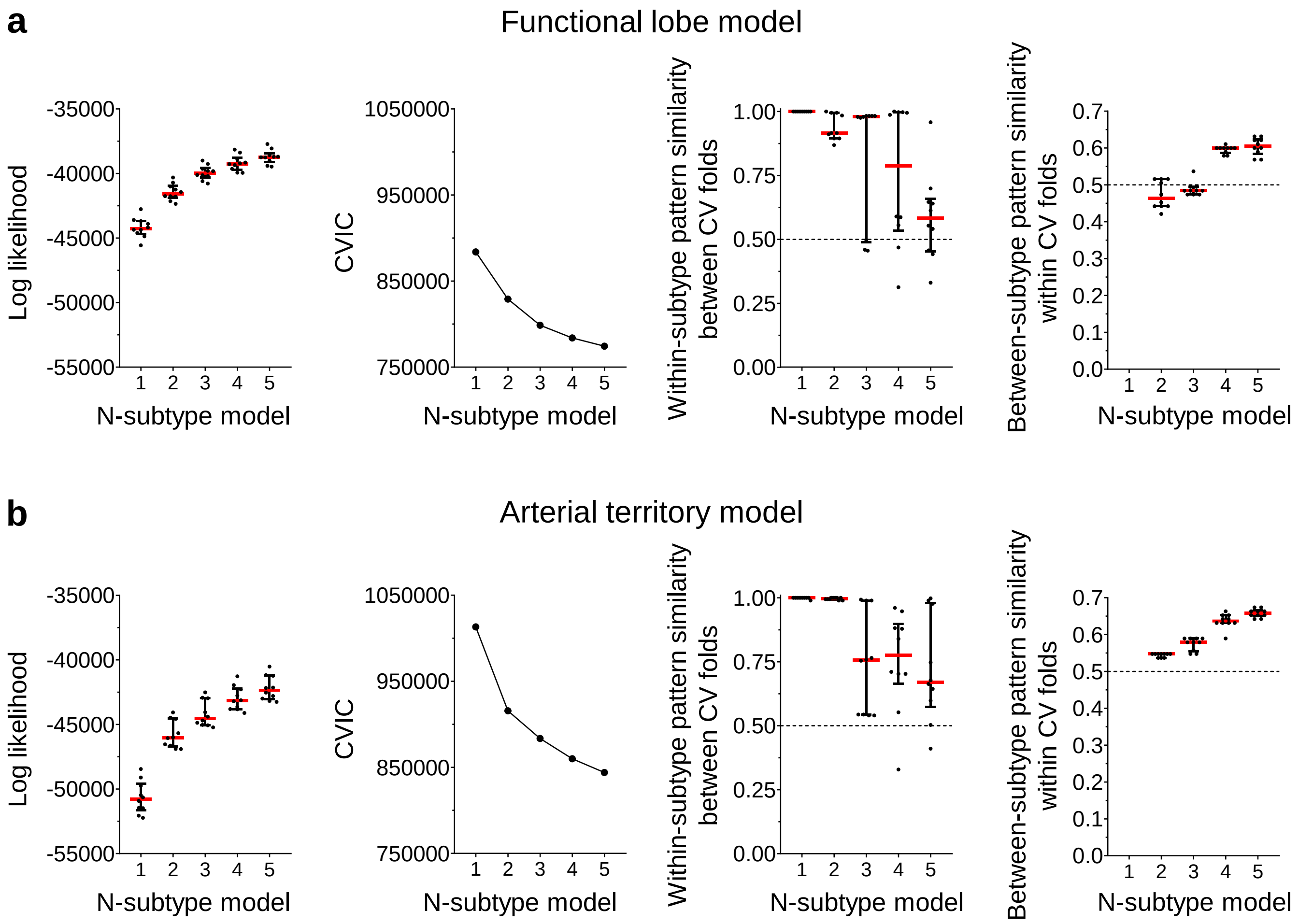

**Supplementary Figure 4 | Voxel-based white matter hyperintensity (WMH) frequency maps across distinct spatiotemporal trajectories of WMH progression in stroke patients.**

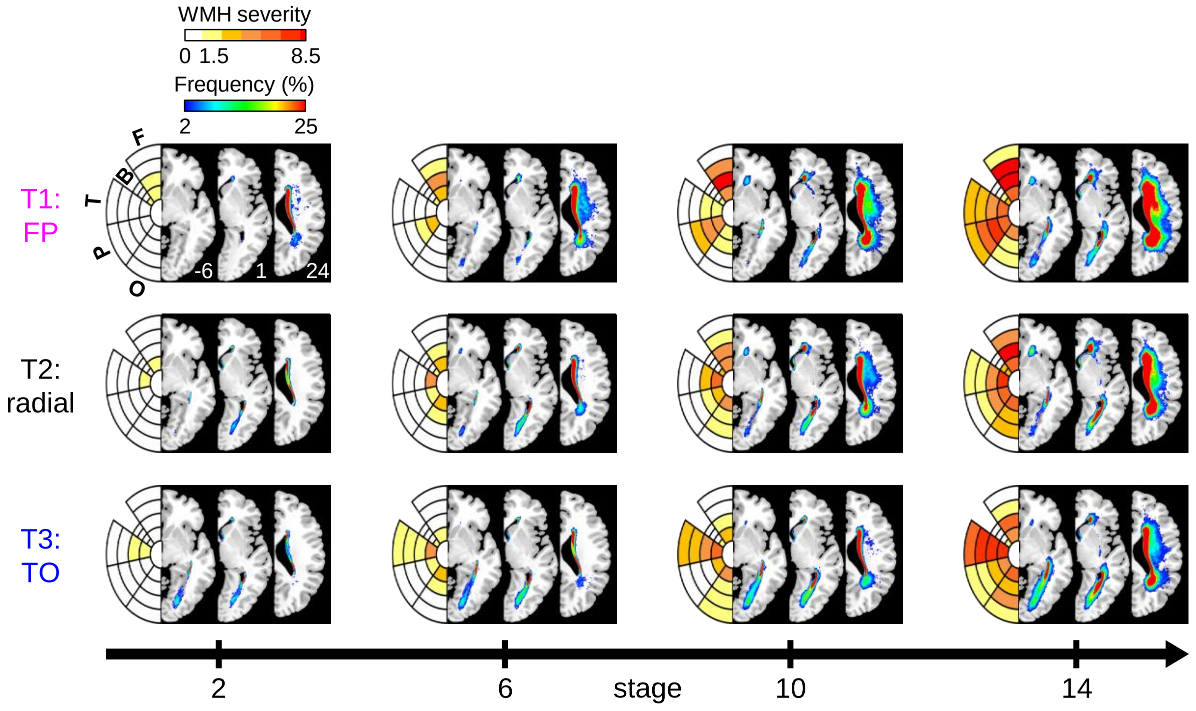

**Supplementary Figure 5 | Distinct demographics, risk factor, and etiology profiles of stroke patients with different white matter hyperintensity (WMH) subtypes without adjustment for age and sex.**

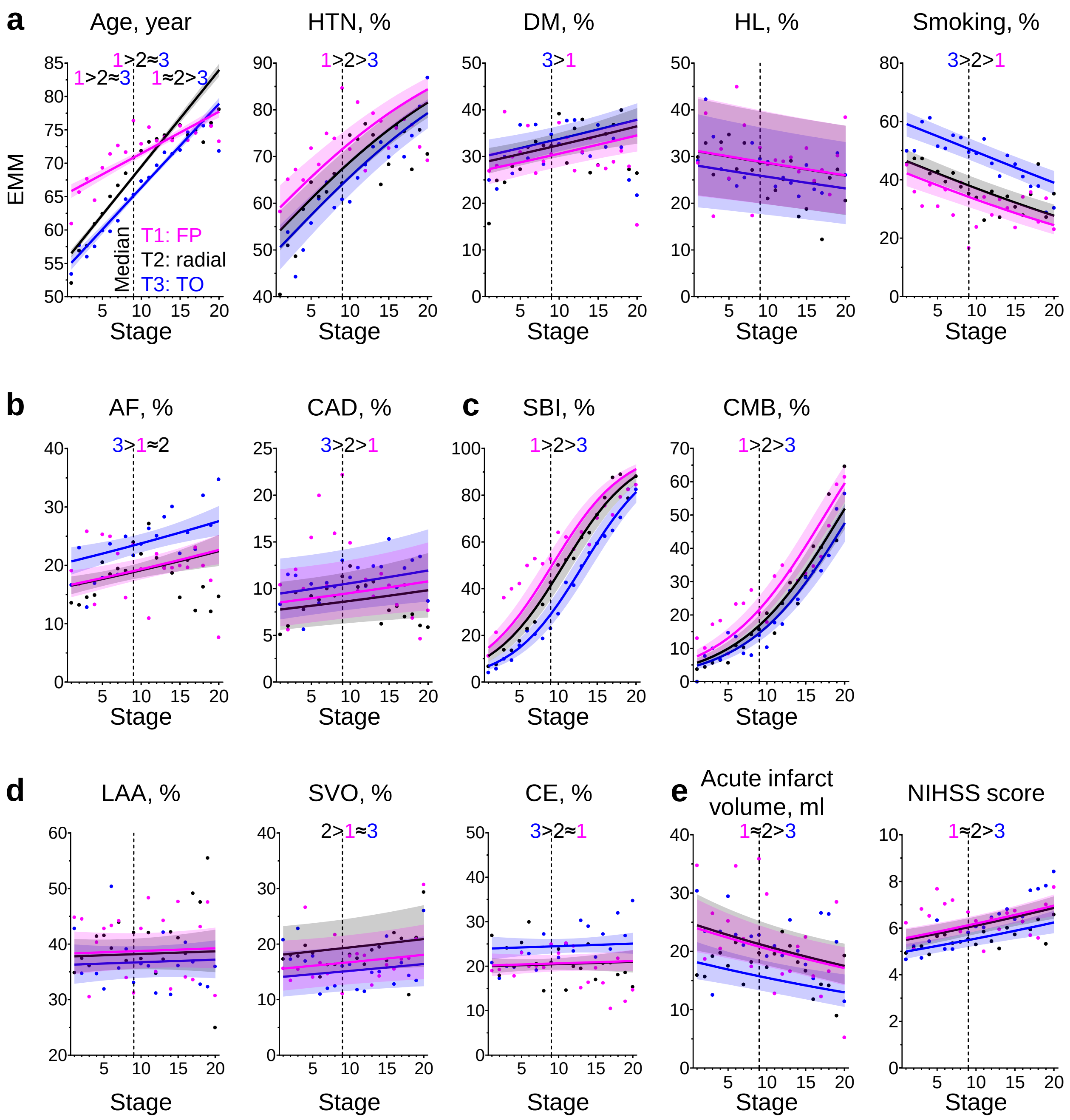

**Supplementary Figure 6 | Proportions of atrial fibrillation (AF) and coronary artery disease (CAD) by subtype and stage of white matter hyperintensity (WMH) progression in high-risk controls.**

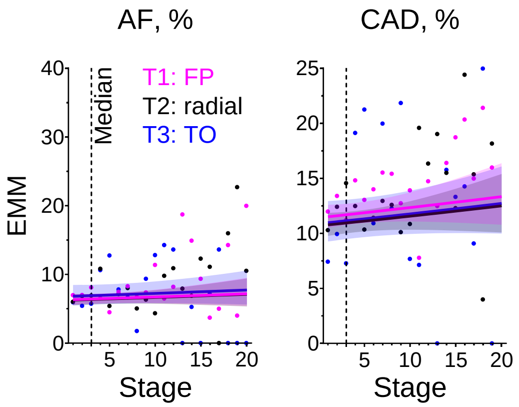

**Supplementary Figure 7 | Comparison of white matter hyperintensity (WMH) stage and volume in discriminating stroke patients from high-risk controls.**

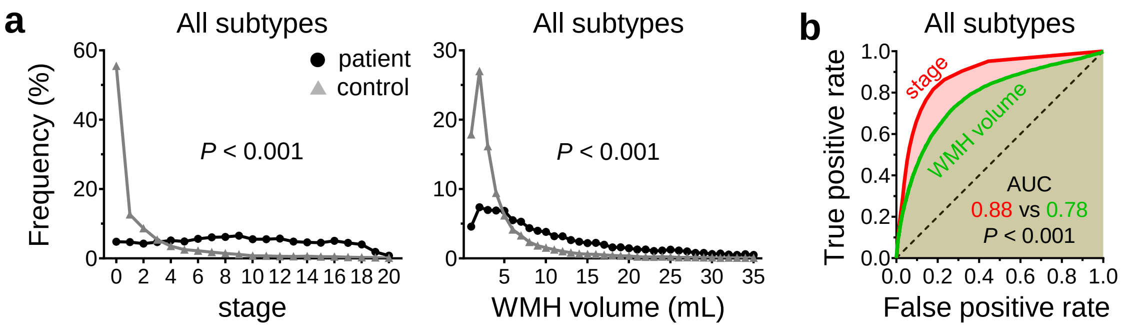

**Supplementary Figure 8 | Bullseye parcellation of cerebral white matter based on arterial territories.**

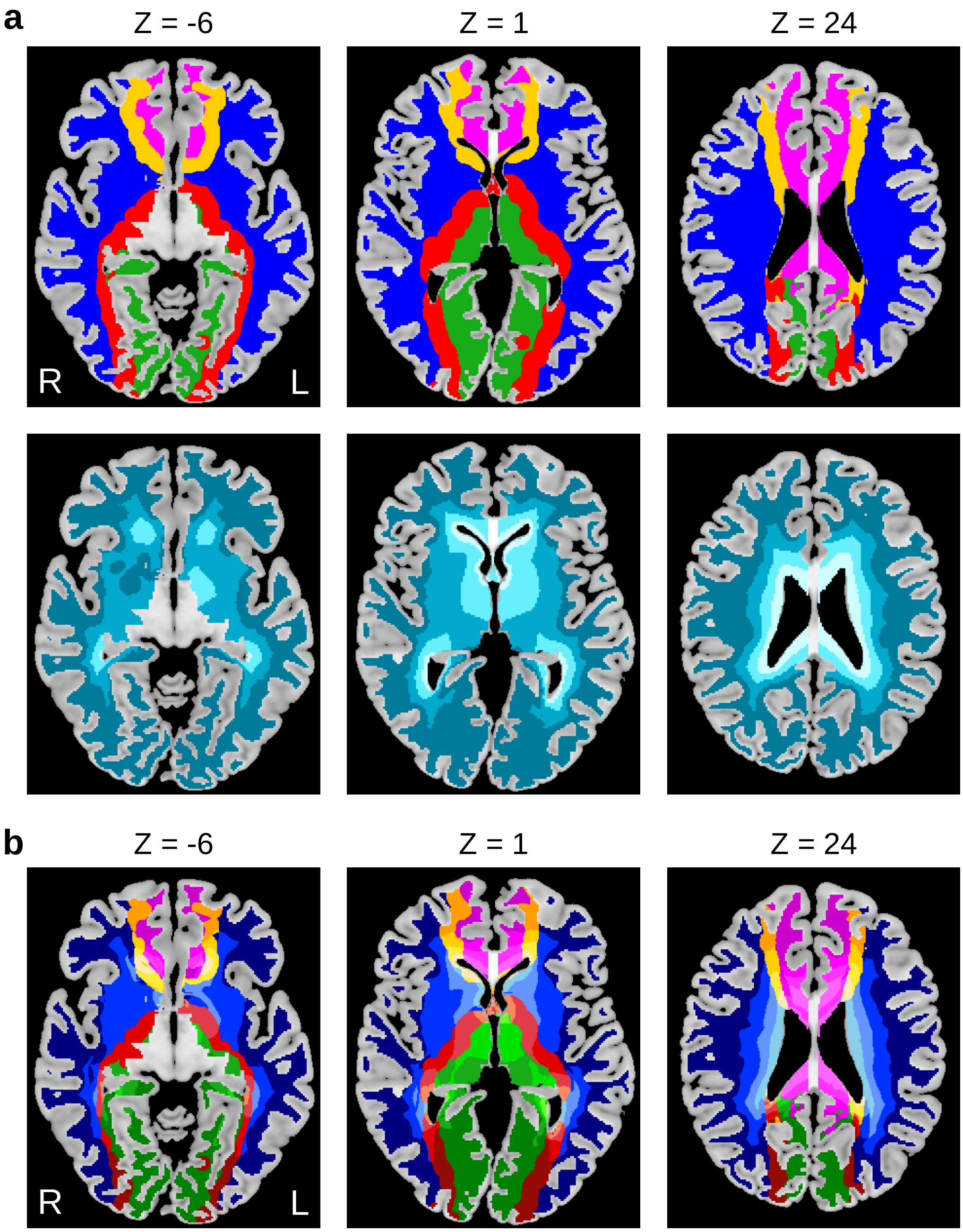

**Supplementary Figure 9 | Distinct spatiotemporal trajectories of white matter hyperintensity (WMH) progression in stroke patients matched to low-risk controls by age and sex**

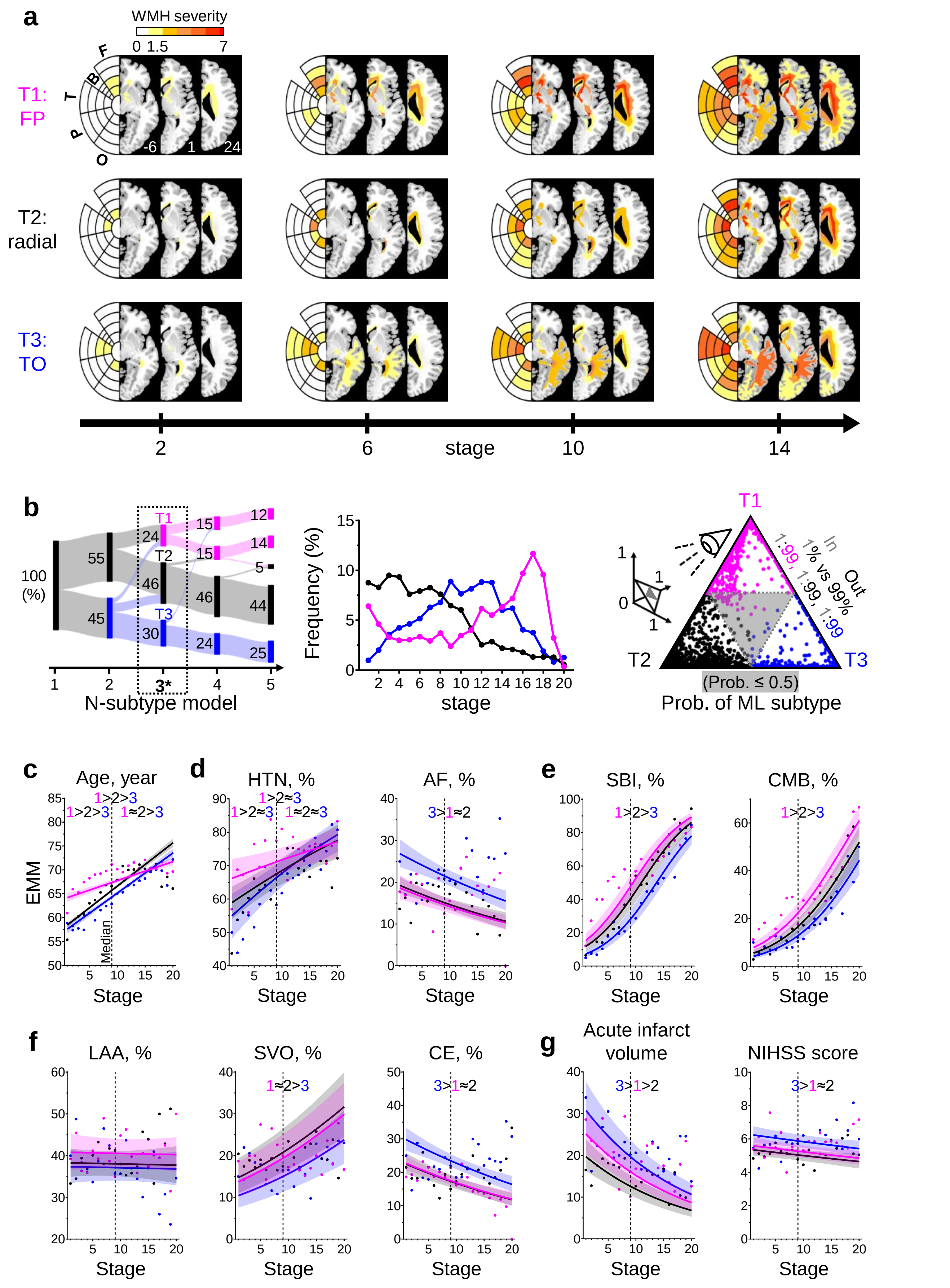

**Supplementary Figure 10 | Post-stroke outcomes by subtype and stage of white matter hyperintensity (WMH) progression in age- and sex-matched stroke patients (to low-risk controls).**

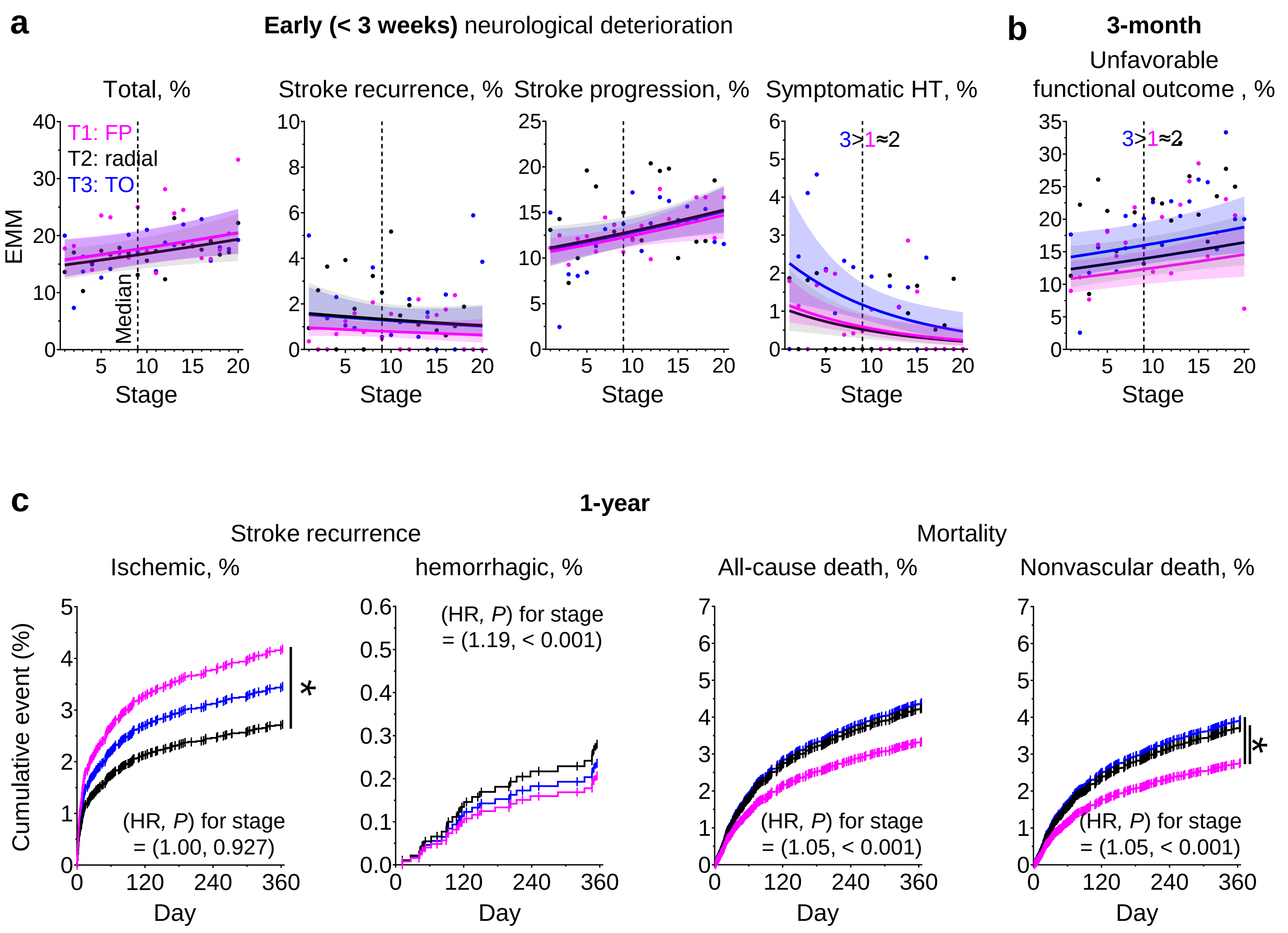
